# Reallocating time from device-measured sleep, sedentary behaviour or light physical activity to moderate-to-vigorous physical activity is associated with lower cardiovascular disease risk

**DOI:** 10.1101/2020.11.10.20227769

**Authors:** Rosemary Walmsley, Shing Chan, Karl Smith-Byrne, Rema Ramakrishnan, Mark Woodward, Kazem Rahimi, Terence Dwyer, Derrick Bennett, Aiden Doherty

## Abstract

**Background:** Moderate-to-vigorous physical activity (MVPA), light physical activity, sedentary behaviour and sleep have all been associated with cardiovascular disease (CVD). Due to challenges in measuring and analysing movement behaviours, there is uncertainty about how the association with incident CVD varies with the time spent in these different movement behaviours.

**Methods:** We developed a machine-learning model (Random Forest smoothed by a Hidden Markov model) to classify sleep, sedentary behaviour, light physical activity and MVPA from accelerometer data. The model was developed using data from a free-living study of 152 participants who wore an Axivity AX3 accelerometer on the wrist while also wearing a camera and completing a time use diary. Participants in UK Biobank, a prospective cohort study, were asked to wear an accelerometer (of the same type) for seven days, and we applied our machine-learning model to classify their movement behaviours. Using Compositional Data Analysis Cox regression, we investigated how reallocating time between movement behaviours was associated with CVD incidence.

**Findings:** We classified accelerometer data as sleep, sedentary behaviour, light physical activity or MVPA with a mean accuracy of 88% (95% CI: 87, 89) and Cohen’s kappa of 0·80 (95% CI: 0·79, 0·82). Among 87,509 UK Biobank participants, there were 3,424 incident CVD events. Reallocating time from any behaviour to MVPA, or reallocating time from sedentary behaviour to any behaviour, was associated with a lower risk of CVD. For example, for a hypothetical average individual, reallocating 20 minutes/day to MVPA from all other behaviours proportionally was associated with 9% (7%, 10%) lower risk of incident CVD, while reallocating 1 hour/day to sedentary behaviour was associated with 5% (3%, 7%) higher risk.

**Interpretation:** Reallocating time from light physical activity, sedentary behaviour or sleep to MVPA, or reallocating time from sedentary behaviour to other behaviours, was associated with lower risk of incident CVD. Accurate classification of movement behaviours using machine-learning and statistical methods to address the compositional nature of movement behaviours enabled these insights. Public health interventions and guidelines should promote reallocating time to MVPA from other behaviours, as well as reallocating time from sedentary behaviour to light physical activity.

**Funding:** Medical Research Council.

## Research in Context

### Evidence before this study

Low levels of moderate-to-vigorous physical activity, low levels of light physical activity, high levels of sedentary behaviour and short and long sleep time have been associated with cardiovascular disease risk. Uncertainty remains about how different combinations of behaviours are associated with risk, in part due to challenges in the measurement (e.g. reliance on self-reported measures or crude device-based measures) and analysis of movement behaviours (e.g. taking appropriate account of behaviours making up the 24 hour day). We searched PubMed for studies published up to the end of September 2020 that investigated the association between device-measured moderate-to-vigorous physical activity, light physical activity, sedentary behaviour or sleep and incident cardiovascular disease in adult populations (search terms in **Supplementary Material**). Four studies were identified, which found that lower levels of sedentary behaviour, higher levels of light physical activity and higher levels of moderate-to-vigorous physical activity were associated with lower cardiovascular disease risk. These studies did not consider sleep and used traditional ‘cut-point’ based methods to identify movement behaviours in device data. Most categorised the exposure and adjusted only partially for time in behaviours other than the behaviour of interest.

### Added value of this study

In this prospective study of 87,509 participants, we used machine-learning methods, developed and validated using a separate free-living dataset, to accurately classify device data as sleep, sedentary behaviour, light physical activity or moderate-to-vigorous physical activity. We used a Compositional Data Analysis approach to investigate how reallocating time between device-measured movement behaviours was associated with cardiovascular disease incidence. We found that, for a hypothetical average individual, reallocating 20 minutes/day to moderate-to-vigorous physical activity from all other behaviours proportionally was associated with 9% (95% CI: 7%, 10%) lower risk of incident cardiovascular disease, whereas reallocating 1 hour/day to sedentary behaviour was associated with 5% (3%, 7%) higher risk.

### Implications of all the available evidence

Emerging methods, such as machine-learning based behaviour classification and Compositional Data Analysis for epidemiological analysis, can provide new health insights. Using these methods we found that reallocating time to moderate-to-vigorous physical activity from other behaviours was associated with lower cardiovascular disease risk and thus should be encouraged. Reallocating time from sedentary behaviour to other behaviours was also associated with lower cardiovascular risk. To strengthen the evidence for causality, intervention studies should examine the health consequences of reallocating time between movement behaviours.

## Introduction

Previous studies have shown low levels of light physical activity^1^ and moderate-to-vigorous physical activity^2^ (MVPA) and high levels of sedentary behaviour^3^ are positively associated with cardiovascular risk, whereas for sleep a U-shaped association has been found.^4,5^ Due to challenges in measuring and analysing movement behaviours, there is uncertainty about how different combinations of movement behaviours are related to cardiovascular disease (CVD) risk.

Recall and reporting bias affect self-reported measurements,^6^ and some behaviours (e.g. light physical activity) are hard to capture.^7^ Device-based measurements address these concerns but introduce new challenges. Many studies use hip-worn devices, where mean wear time is typically <15 hours/day and sleep is not measured.^8^ Behaviours have typically been classified using ‘cut-point’ based methods, which use an acceleration threshold to distinguish behaviours.^9^ These methods can only distinguish a limited set of behaviours and are prone to substantial misclassification.^9–11^ Emerging machine-learning methods could allow a wider range of behaviours to be classified accurately,^9–11^ providing they are developed and validated using free-living datasets (rather than laboratory-based data).^9–11^

There is uncertainty about how movement behaviours are associated with CVD, as analyses often neglect the fact that behaviours occur alongside one another (e.g. individuals spending large amounts of time sedentary may also typically spend small amounts of time in light physical activity). In analyses where movement behaviours are not mutually adjusted for, residual confounding is likely. Further complicating this, a person who increases time spent in one behaviour must compensate by decreasing time spent in others. This means that analyses should address the effect of reallocating time between behaviours (‘isotemporal substitution’).^12^ It also means that movement behaviour data is compositional data, whereby only the relative time spent in different behaviours (and not the absolute time in each behaviour) is informative.^13,14^ An individual cannot increase time spent in light physical activity while holding time in other behaviours fixed. However, they can increase time spent in light physical activity relative to other behaviours, while holding each of those behaviours fixed as a proportion of the remaining day. Methods for analysing compositional data aim to capture and model the relative values of variables.^13,14^

The objective of this study was to investigate the association between device-measured movement behaviours and risk of incident CVD in middle-to older-aged adults by:

(i) Using free-living ‘ground truth’ data to develop and validate a machine-learning model to classify movement behaviours from wrist-worn accelerometer data.

(ii) Applying this new model to classify movement behaviours of 87,509 UK Biobank participants who wore an accelerometer.

(iii) Characterising the association between device-measured movement behaviours and incident CVD, accounting for the compositional nature of movement behaviours.

## Methods

### UK Biobank: a large prospective cohort study

UK Biobank is a population-based prospective cohort study of over 500,000 participants in England, Scotland and Wales (protocol available at https://www.ukbiobank.ac.uk/key-documents/). Between 2006 and 2010, individuals aged 40-69 living within roughly 25 miles of an assessment centre were recruited by letter (all eligible individuals were identified from National Health Service records; response rate 5.5%).^15^ At baseline, participants attended an assessment involving a touchscreen questionnaire, biological sampling, an interview by a trained interviewer, and anthropometric measurements.^15^ UK Biobank received ethical approval from the National Health Service National Research Ethics Service (Ref 11/NW/0382).

### Device-based measures of movement behaviours in UK Biobank

Between June 2013 and December 2015, participants with a valid email address (excluding North West region due to participant burden concerns) were invited to wear an accelerometer. 106,053 consenting participants were sent an Axivity AX3 wrist-worn triaxial accelerometer to be worn on the dominant wrist for seven days.^16^ A readable accelerometer dataset was obtained from 103,684 participants. Initial data processing followed established methods:^16^ participants were excluded if the device could not be calibrated, if they had less than three days of data or did not have data in each one hour period of the 24 hour cycle (with non-wear time defined as unbroken episodes of at least 60 minutes during which standard deviation of each axis of acceleration was less than 13.0 mg^16^), if more than 1% of readings were ‘clipped’ (fell outside the device’s dynamic range of ±8g) before or after calibration, or if the average acceleration was implausibly high (>100mg).^16^ Recording interruptions and non-wear time were imputed as the mean behaviour in the corresponding minute of the day on remaining days.

### Classification of movement behaviours using machine-learning methods

#### CAPTURE-24

CAPTURE-24, an accelerometer validation study of 152 adults aged 18-91 recruited by advertisements in Oxford, UK, in 2014-2015,^11^ was used to develop machine-learning classification methods. Participants were asked to wear an Axivity AX3 wrist-worn accelerometer for 24 hours, wear a Vicon Autographer wearable camera while awake during that period, and keep a time use diary.^10^ Using camera images and time use diaries, trained annotators annotated accelerometer data with labels from the Compendium of Physical Activities.^17^ Fine-grained labels were mapped to sleep, sedentary behaviour, light physical activity, and MVPA (see **Supplementary Material** and **Table S1**). Describing intensity in METs (Metabolic Equivalent of Task), which measure energy expenditure relative to energy expenditure in quiet sitting, these behaviours were defined as:

1. Sleep: non-waking behaviour.
2. Sedentary behaviour: waking behaviour at <1·5 METs in a sitting, lying or reclining posture.^18^
3. Light physical activity: waking behaviour at <3 METs not meeting the sedentary behaviour definition.
4. MVPA: all behaviour at ≥3 METs.^17^

CAPTURE-24 received ethical approval from the University of Oxford Inter-Divisional Research Ethics Committee (Ref SSD/CUREC1A/13-262).

#### Machine-learning for behaviour classification

Using this labelled data from the CAPTURE-24 study, a balanced Random Forest with 100 decision trees was trained to classify the behaviour in 30-second time windows using 50 rotation-invariant time and frequency domain features of the accelerometer signal (**Table S2**). As the Random Forest did not use time sequence information, the behaviour sequence was smoothed using a Hidden Markov model. This model treated the Random-Forest-predicted behaviours as ‘emissions’ from an underlying true behaviour sequence, and used the Viterbi algorithm to identify the most likely underlying true sequence given the observed sequence^19^. Transition probabilities between different behaviours were determined using camera validation data, and probability of the Random Forest predicting each behaviour conditional on the true behaviour was estimated using out-of-bag estimates from the Random Forest. This model structure closely followed our previous work,^10,11^ and more detail is given in the **Supplementary Material**.

Performance was evaluated using Leave-One-Participant-Out Cross-Validation. Accuracy was used to assess overall agreement between annotator-assigned ‘ground truth’ labels and model-assigned labels, and Cohen’s kappa was used to assess agreement beyond that expected by chance. Precision and recall were used to assess performance on each behaviour, and the confusion matrix was used to show classification patterns for examples of each behaviour. Accuracy, Cohen’s kappa, precision and recall were calculated for each participant individually, and we computed their mean (across participants). To examine how sensitive mean precision and recall were to the results of participants with few examples of a behaviour, the mean was recalculated excluding participants with up to 20 minutes of a particular behaviour (the **Supplementary Material** contains more detail on performance evaluation). Face validity of the behaviour classification method applied to UK Biobank data was assessed by plotting the behaviour profile of UK Biobank participants across the day.

### Ascertainment of cardiovascular disease endpoints

UK Biobank has ongoing passive follow-up via linkage to Hospital Episode Statistics (HES; hospital diagnoses from the National Health Service, the provider of almost all UK healthcare), and the UK death register.^15^ Cardiovascular disease was defined as ICD-10 codes I20-25 (ischaemic heart diseases) or I60-69 (cerebrovascular diseases) appearing in HES or on the death register. Participants with cardiovascular disease prior to accelerometer wear, either HES-recorded or self-reported in the baseline questionnaire, were excluded. Participants who did not experience a cardiovascular disease outcome were censored at death or the end of the study period as appropriate (30/06/2020 for participants in England, 29/02/2016 for participants in Wales and 31/10/2016 for participants in Scotland).

### Compositional Data Analysis for movement behaviour data

A Compositional Data Analysis approach was used in the statistical analyses. This approach uses log-ratios (log-transformed ratios between movement behaviours) to describe and adjust for the movement behaviour composition. By using ratios between behaviours, the relative time in different behaviours, rather than the absolute time in any given behaviour, is modelled. For this analysis, we used isometric log-ratio pivot coordinates, a particular set of log-ratios which is widely used in movement behaviour research (see **Supplementary Material** for more detail).^14,20^

Our results are described by isotemporal substitution plots, which show the hazard ratio associated with reallocating time from one behaviour to another behaviour, and by the hazard ratio associated with particular reallocations of time between behaviours (e.g. reallocating 1 hour/day to sedentary behaviour from all other behaviours proportionally).^21^ These hazard ratios are relative to the mean behaviour composition among included participants, so can be interpreted as showing the outcome associated with reallocating time between behaviours for a hypothetical average individual.

### Statistical analyses

Multivariable-adjusted Cox proportional hazards regression models, with age as the timescale, were used to investigate the association between the movement behaviour composition and incident cardiovascular disease. To address potential sources of confounding, the main analysis was stratified by sex and adjusted for ethnicity (Asian, Black, Other, White), smoking status (current, ex-or never smoker), frequency of alcohol consumption (never, <3 times/week, 3+ times/week), fresh fruit and vegetable consumption (<3, 3-4.9, 5-7.9, 8+ servings/day), frequency of red and processed meat consumption (<1, 1-1·9, 2-3.9, 4+ times/week), frequency of oily fish consumption (<1, 1, 2-4, >4 times/week), education (school leaver, further education, higher education) and deprivation (quarter of Townsend Deprivation Index in the study population). A minimally adjusted analysis was stratified by sex and used age as the timescale but had no further adjustment for potential confounders. To investigate the impact of BMI an additional analysis was adjusted for BMI as a continuous variable. A further multivariable-adjusted analysis was performed with fatal cardiovascular events as the outcome. All adjustment variables were measured at baseline assessment (**Table S3** gives more details on all variables used in the analysis), and variables were not adjusted for if they were likely mediators of the association between movement behaviours and cardiovascular disease.

Participants with missing data in any adjustment variable were excluded. The proportional hazards assumption was tested component-wise and globally using the Grambsch-Therneau test with the Kaplan-Meier transformation,^20^ and there was no evidence (at the 5% level) that it was violated in the main analysis. Plots of the Schoenfeld residuals were also examined. Results were reported according to STROBE guidelines,^22^ and all confidence intervals are 95% confidence intervals. Software is described in **Supplementary Material**.

### Sensitivity analyses

The impact of reverse causality was assessed first by excluding the initial two years of follow-up and any events within it. A further analysis additionally excluded participants who self-reported poor health or use of diabetes or cardiovascular disease-related medications at baseline or who had a prior hospital admission for any condition of the circulatory system (I00-I99 as a primary diagnosis e.g. admission for heart failure or aortic aneurysm).

To investigate unmeasured and residual confounding we used a negative control outcome of accidents without a plausible mechanistic link to movement behaviours (accidents excluding falls, cycling accidents and intentional self-harm; see **Table S3**).^23^ We also used E-values to assess the minimum strength of association that an unmeasured confounder would need with both exposure and outcome to explain away the observed association (see **Supplementary Material**).^24,25^

## Results

### Movement behaviour classification in the training dataset

Our machine-learning method accurately classified movement behaviours in accelerometer data: when evaluated using Leave-One-Participant-Out Cross-Validation in the CAPTURE-24 study (**Table S4**), mean per-participant accuracy was 88% (95% CI: 87, 89) and mean per-participant Cohen’s kappa was 0·80 (95% CI: 0·79, 0·82). Mean per-participant precision and recall for each behaviour show most examples of all behaviours were correctly classified, with highest performance for sleep (**Figure S1**). Misclassifications were most common between similar behaviours (**Table S5**). As expected, classification performance was worse on individuals with very few true examples of a behaviour (**Figure S1**). The behaviour classification showed high face validity when applied to UK Biobank participants’ data (**Figure S2**).

### Analyses in the UK Biobank

#### Baseline characteristics

After excluding participants with poor quality accelerometer data, participants with prevalent disease and participants with missing data, 87,509 UK Biobank participants were included in the Cox regression analysis for incident cardiovascular disease (**Figure 1**).

**Figure 1:**
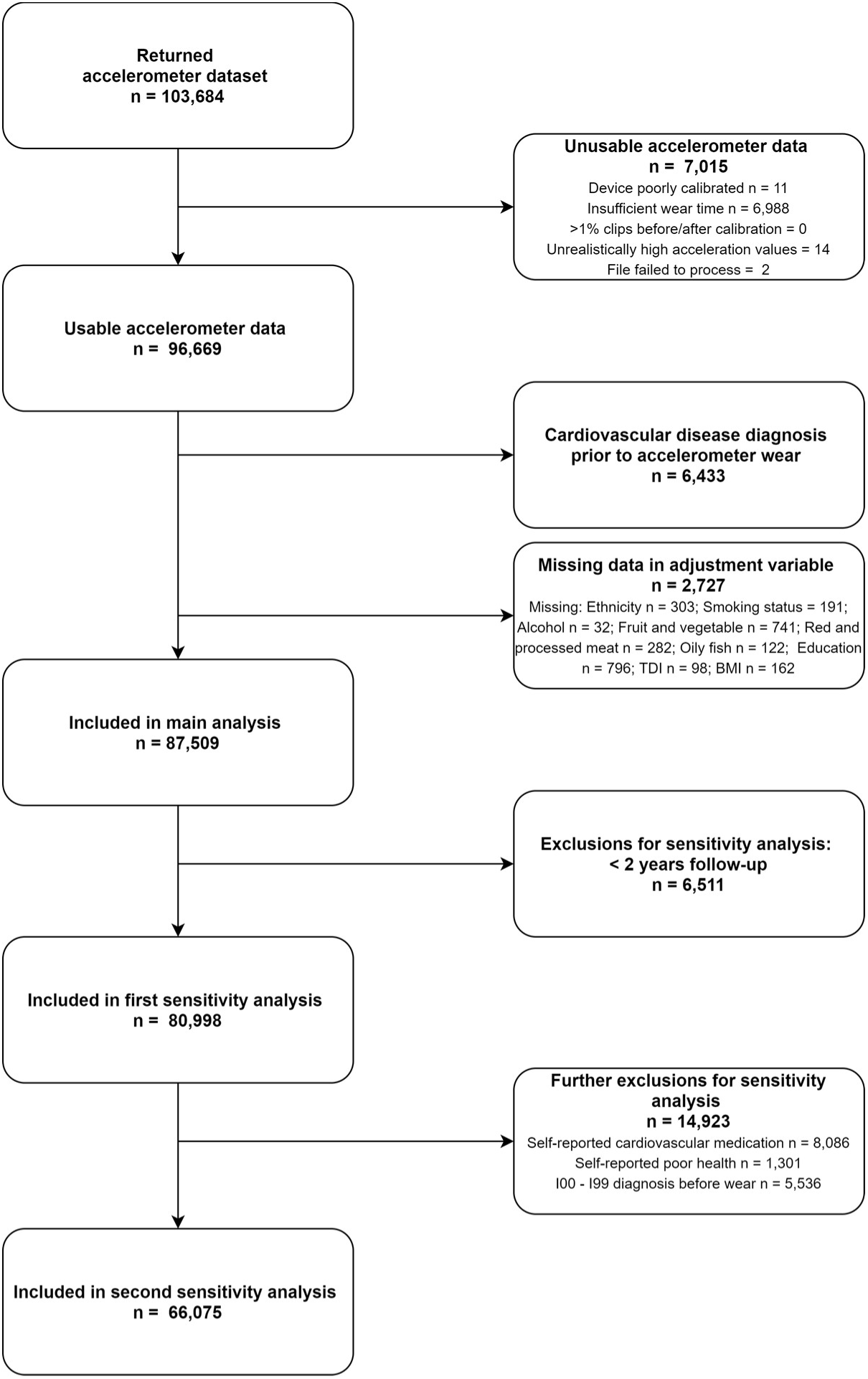
Participant flow diagram for the analysis of movement behaviours and incident cardiovascular disease in UK Biobank participants.

The mean composition of movement behaviours (the daily movement behaviours of a hypothetical average individual) was 8.8 hours/day sleep, 9.3 hours/day sedentary behaviour, 5.6 hours/day light physical activity and 21 minutes/day MVPA (**Figure 2**).

**Figure 2:**
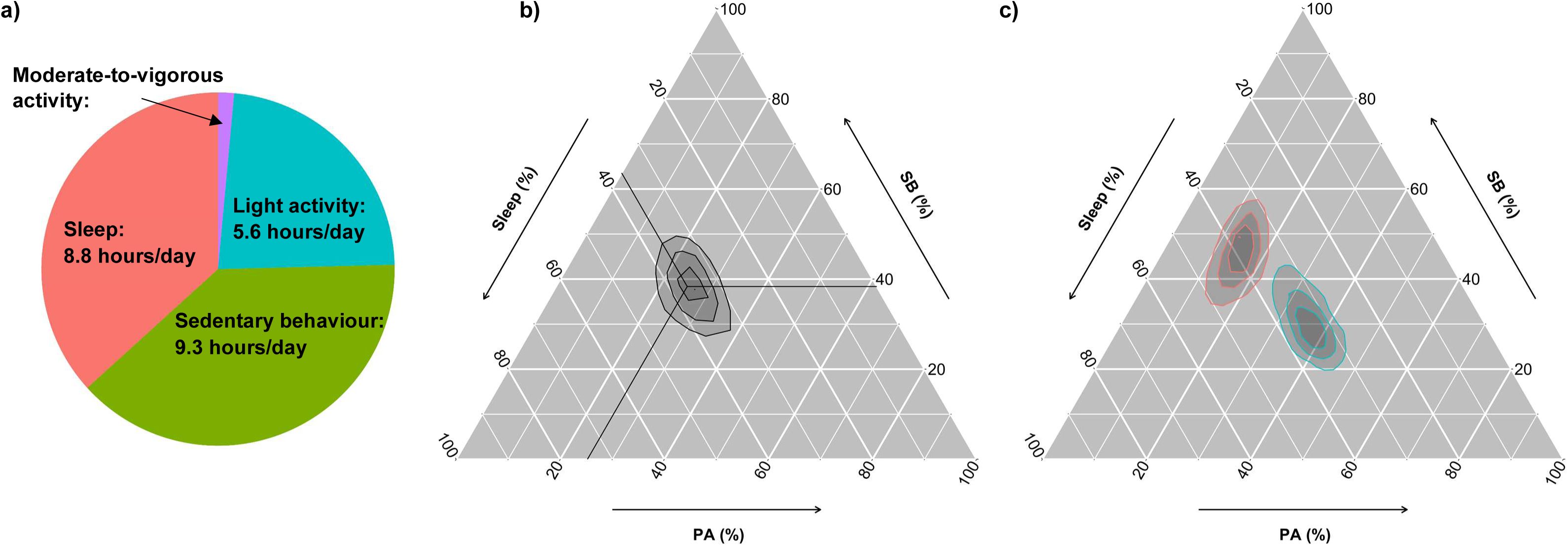
Distribution of movement behaviours in 87,509 UK Biobank participants. a) Mean movement behaviour composition among UK Biobank participants. b) Movement behaviours of UK Biobank participants on a ternary density plot, showing sleep, sedentary behaviour (SB) and physical activity (PA; combines light and moderate-to-vigorous physical activity). The crosshair marks the compositional mean. 25% of participants fall between each pair of lines. The behaviour composition at a point can be found by tracing out (parallel to the white lines) from the point to the axes. c) Ternary density plot showing the behaviour distribution of the 5% most active (blue) and 5% least active (red) UK Biobank participants by average acceleration. 25% of participants in each group fall between each pair of lines.

When considering movement behaviours according to participant characteristics, notable differences included that women had higher levels of light physical activity than men, and lower sedentary time and MVPA (**Table 1**). Older participants spent less time in MVPA than younger participants (**Table 1**). Participants with higher BMI spent less time in light physical activity and MVPA than participants with lower BMI, and spent more time sedentary (**Table 1**).

**Table 1:**
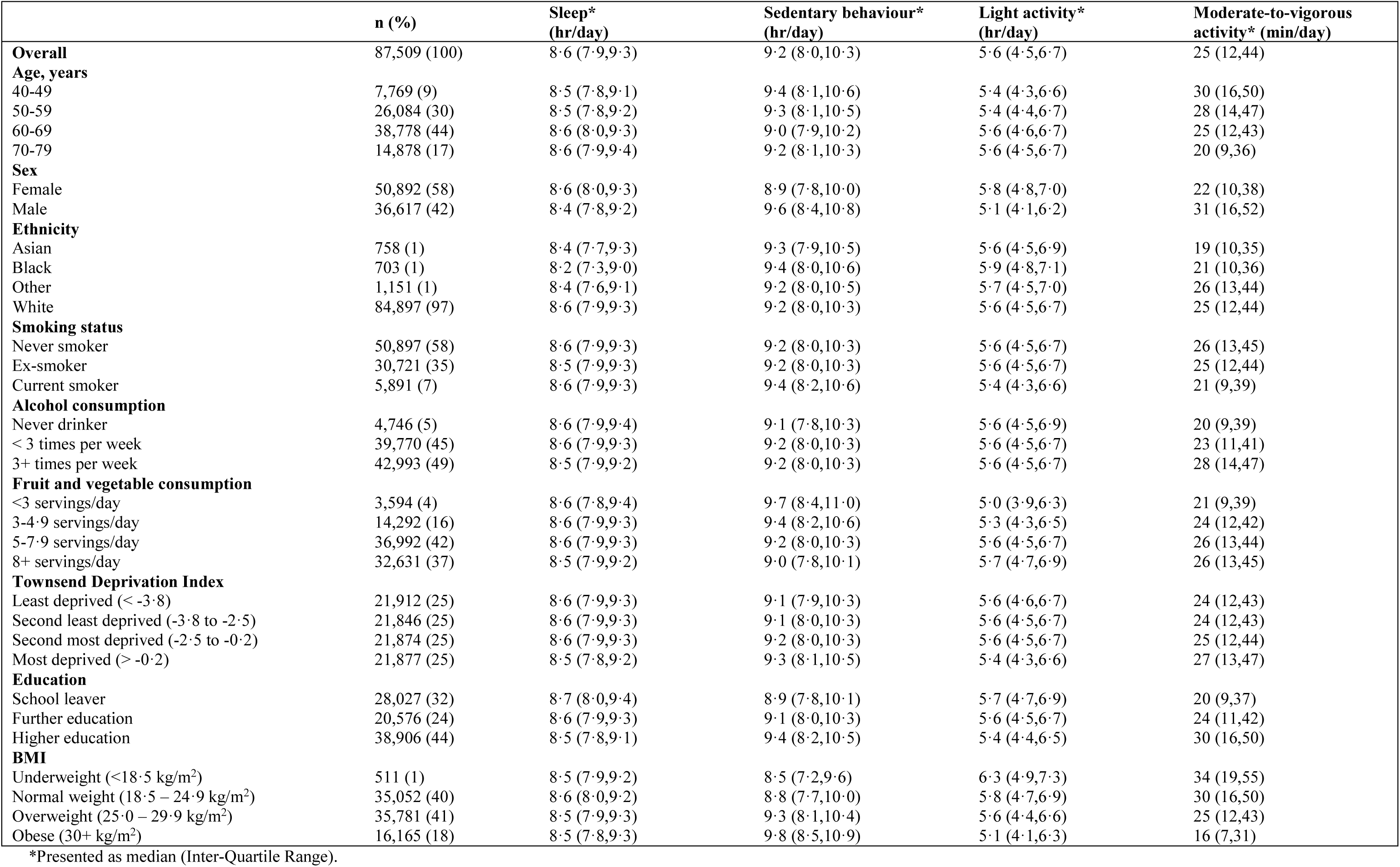
Movement behaviours of 87,509 UK Biobank participants by participant characteristics.

#### Associations with incident cardiovascular disease

Over 445,752 person years of follow-up (median 5.4 years, maximum 7.1 years), there were 3,424 incident cardiovascular disease events. Reallocating time from sedentary behaviour to light physical activity was associated with a lower risk of cardiovascular disease (**Figure 3a**): for a hypothetical average individual, the hazard ratio (HR) associated with reallocating 1 hour/day from light physical activity to sedentary behaviour was 1·04 (95% CI: 1·01, 1·06), while the HR associated with reallocating 1 hour/day from sedentary behaviour to light physical activity was 0·96 (0·94, 0·98). Reallocating time from sedentary behaviour to MVPA was associated with more pronounced lower risk of cardiovascular disease (**Figure 3b**): for an average individual, the HR associated with reallocating 15 minutes/day from MVPA to sedentary behaviour was 1·19 (1·16, 1·23), while the HR associated with reallocating 15 minutes/day from sedentary behaviour to MVPA was 0·92 (0·91, 0·94). Reallocating time from light physical activity to MVPA was also associated with a lower risk of cardiovascular disease (**Figure 3c**).

**Figure 3:**
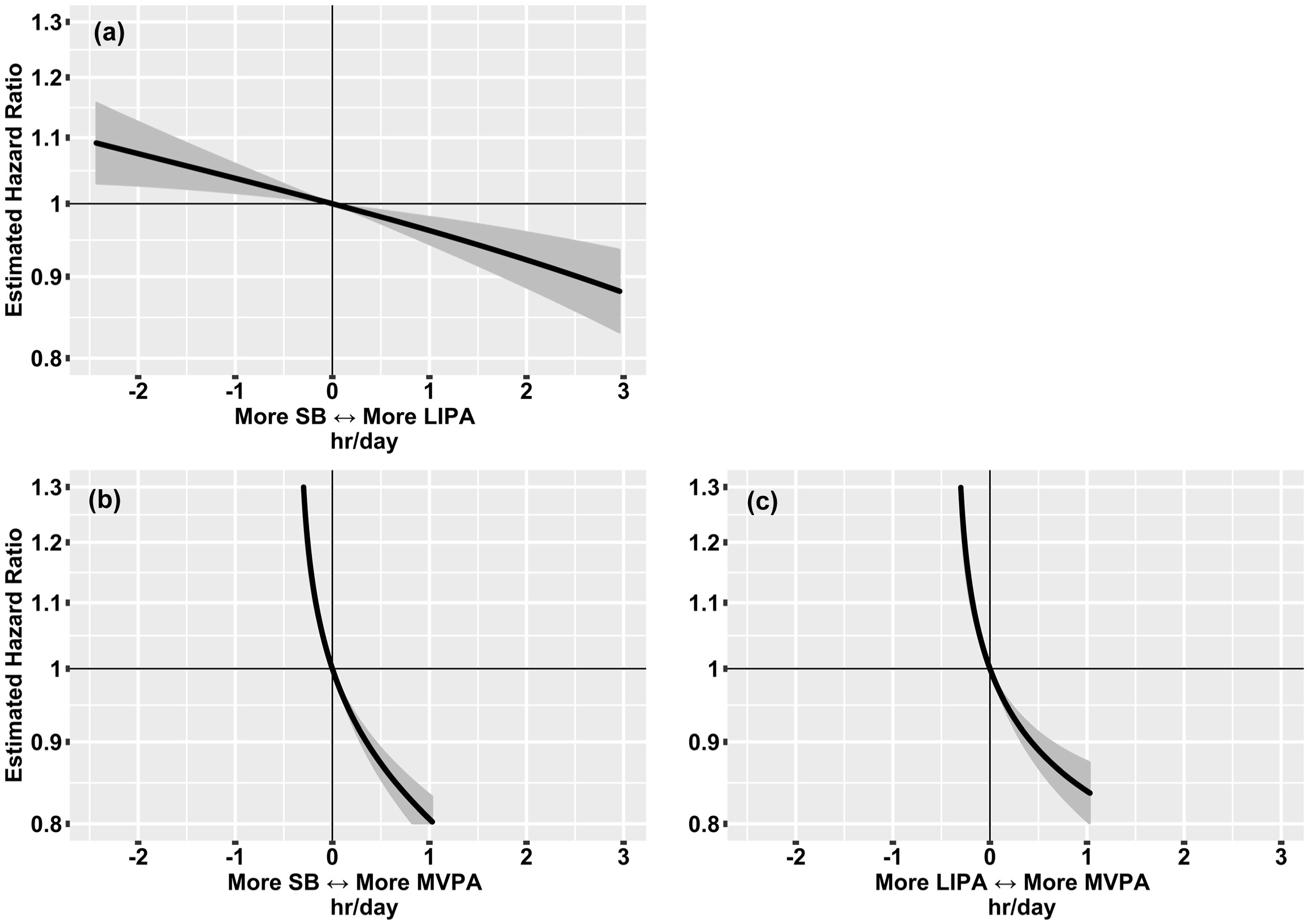
Hazard ratios for incident cardiovascular disease associated with balance between movement behaviours in 87,509 UK Biobank participants.* (a) sedentary behaviour (SB) and light physical activity (LIPA) (b) sedentary behaviour and moderate-to-vigorous physical activity (MVPA) and (c) light physical activity and moderate-to*-*vigorous physical activity. *Model based on 3,424 events in 87,509 participants. All relative to the mean behaviour composition (8·8 hours/day sleep, 9·3 hours/day sedentary behaviour, 5·6 hours/day light physical activity, 0·35 hours/day (21 minutes/day) moderate-to-vigorous physical activity). Model used age as the timescale, was stratified by sex and was additionally adjusted for ethnicity, smoking status, alcohol consumption, fresh fruit and vegetable consumption, red and processed meat consumption, oily fish consumption, deprivation and education.

We found that, for an average individual, reallocating 20 minutes/day to MVPA from all other behaviours proportionally was associated with 9% (7%, 10%) lower risk of cardiovascular disease (**Figure 4**; 28% of the study population exceeded this level of MVPA). Reallocating 1 hour/day to sedentary behaviour, from all other behaviours proportionally, was associated with 5% (3%, 7%) higher risk of cardiovascular disease (**Figure 4;** 26% of the study population exceeded this level of sedentary behaviour). Reallocating 1 hour/day to light physical activity, from all other behaviours proportionally, was not significantly associated with cardiovascular disease risk (**Figure 4**).

**Figure 4:**
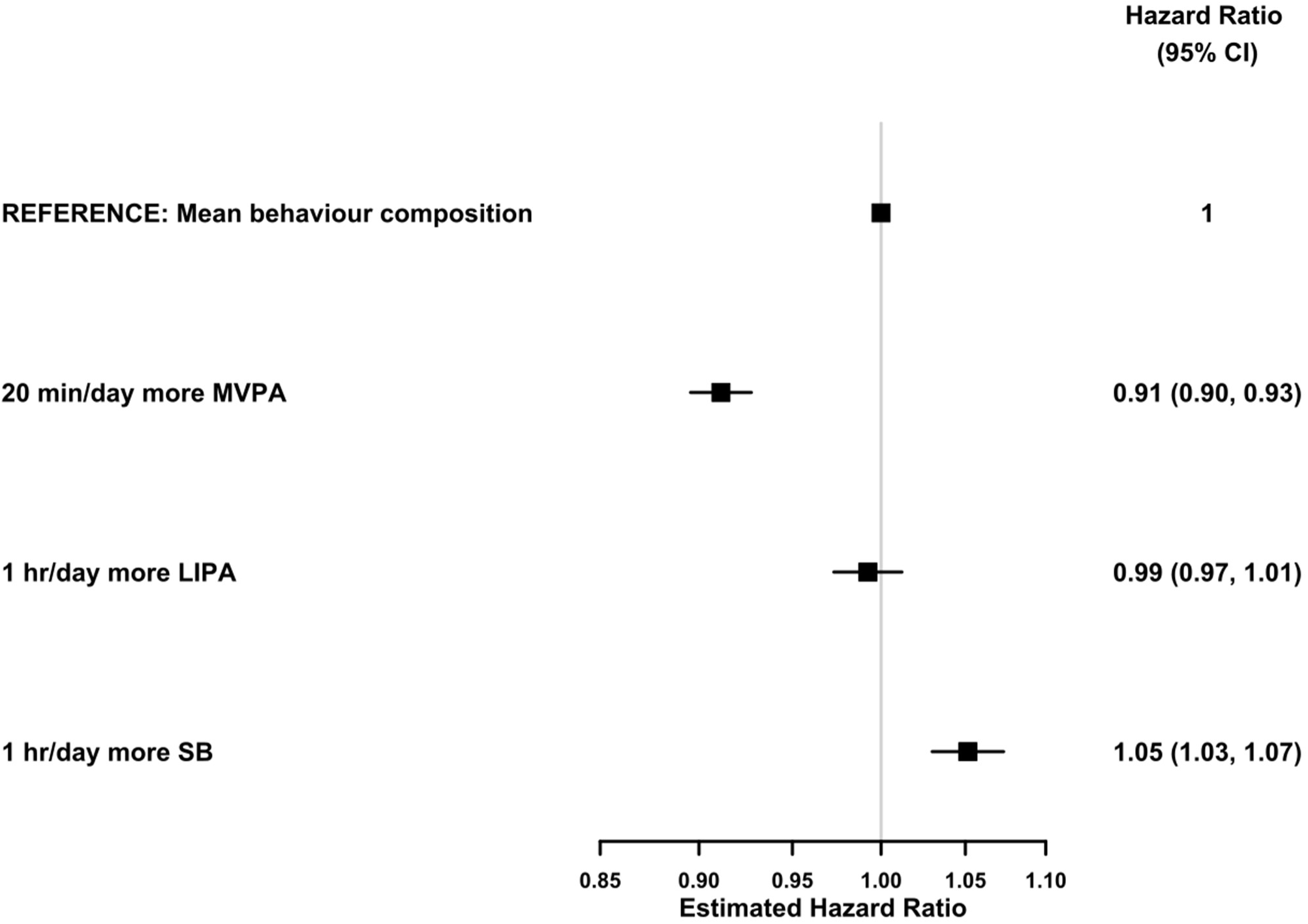
Hazard ratios for incident cardiovascular disease associated with reallocating time to named behaviour, from all other behaviours proportionally, in 87,509 UK Biobank participants.* *Model based on 3,424 events in 87,509 participants. All relative to the mean behaviour composition (8·8 hours/day sleep, 9·3 hours/day sedentary behaviour, 5·6 hours/day light physical activity, 0·35 hours/day (21 minutes/day) moderate-to-vigorous physical activity) and more time in named behaviour reallocated from all other behaviours proportionally. Model used age as the timescale, was stratified by sex and was additionally adjusted for ethnicity, smoking status, alcohol consumption, fresh fruit and vegetable consumption, red and processed meat consumption, oily fish consumption, deprivation and education.

Results from the multivariable-adjusted model were only slightly attenuated compared to those from a minimally adjusted model (**Figure S4**). Further adjustment for BMI showed that there was some attenuation of the association between movement behaviours and incident cardiovascular disease (**Figure S5**). For example, the 9% (7%, 10%) lower risk of cardiovascular disease associated with reallocating 20 minutes/day to MVPA, was reduced to a 7% (5%, 9%) lower risk after adjustment for BMI. Associations for fatal cardiovascular events were similar to those for all cardiovascular events, with some suggestion that reallocating time from sedentary behaviour to light physical activity was more strongly associated with fatal events (**Figure S6**).

Removing the first two years of follow-up attenuated all associations only minimally (**Figure S7**). Further restricting to a healthy subgroup had only a small effect on most associations, but the association for reallocating time from sedentary behaviour to light physical activity was substantially attenuated (**Figure S7**).

Analyses suggested residual and unmeasured confounding had a modest impact on the main findings. Specifically, some movement behaviours were associated with the negative control outcome, suggesting a small impact of residual confounding (**Figure S8**). The E-values indicated that a substantial degree of unmeasured confounding would be required to reduce the observed associations to the null (**Figure S9**). For example, the E-value of 1·42 (for reallocating 20 minutes/day to MVPA, from all other behaviours) shows an unmeasured confounder would need to be associated with at least a 1·42-fold increase in risk for both exposure and outcome to explain away the observed association.

## Discussion

We found that reallocating time to MVPA from sleep, sedentary behaviour or light physical activity or reallocating time from sedentary behaviour to light physical activity or sleep was associated with lower risk of cardiovascular disease. Per minute, the most pronounced differences in risk were seen for MVPA. For a hypothetical average individual, reallocating 20 minutes/day to MVPA from all other behaviours proportionally was associated with 9% (7%, 10%) lower risk of cardiovascular disease. In contrast, reallocating 1 hour/day to sedentary behaviour, from all other behaviours proportionally, was associated with 5% (3%, 7%) higher risk. BMI explained a modest proportion of the association between movement behaviours and incident cardiovascular disease.

Our findings extend previously reported results by showing how reallocating time between behaviours is associated with cardiovascular risk (after adjustment for other behaviours). The results of this study are consistent with our previous results, which showed a dose-response association across quartiles of device-measured moderate physical activity for cardiovascular events in the UK Biobank (with 54% lower risk in the highest quartile for moderate physical activity compared to the lowest).^26^ They are also consistent with results from a study of community-dwelling older women in the US, which found 69% higher risk of incident cardiovascular disease in the highest quartile for device-measured sedentary behaviour compared to the lowest,^27^ and 22% lower risk of incident cardiovascular disease in the highest quartile for light physical activity compared to the lowest.^11^ While we are not aware of previous applications of a Compositional Data Analysis approach in the context of incident cardiovascular disease, McGregor et al investigated the association between movement behaviours and risk of death among 1,468 US adults (135 deaths) and found associations similar to our findings for cardiovascular disease mortality.^28^

Our machine-learning methods accurately identified movement behaviours in accelerometer data, achieving 88% (95% CI: 87, 89) accuracy and Cohen’s kappa of 0·80 (0·79, 0·82) in free-living data. This represents an improvement on previously reported machine-learning approaches (Cohen’s kappa 0·80 vs 0·68), likely due to careful curation of the behaviour classes in labelled data by two reviewers.^11^ While evidence from free-living data is scarce, our approach had better performance than has been reported for traditional ‘cut-point’ approaches.^9^ However, although comparable with UK estimates from other sources,^29^ sleep measurements should be interpreted cautiously: ‘ground truth’ labels for sleep came from a time use diary, which may identify time in bed rather than physiological sleep.

This study has several strengths, notably including the use of device-based measurements to characterise movement behaviours in a large, comprehensive prospective study. Compared to self-reported measurements of behaviour, device-based measurements are at reduced risk of recall and reporting bias,^6^ and they can capture behaviours such as light physical activity well.^7^ The use of a wrist-worn device with a full 24-hour wear protocol (with high compliance) allowed the full day of behaviours to be measured.^16^ The use of free-living data with ‘ground truth’ behaviour labels to develop and validate behaviour classification methods ensures they perform well in real-world settings. All methods used in this study are open-source and available for use in other wrist-worn accelerometer datasets. A major strength of the analysis in this study is the appropriate modelling of 24-hour behaviours using a Compositional Data Analysis approach.^13,14^

An important limitation of any observational study is the possibility of reverse causality bias.^30^ After removing the first two years of follow-up, associations were only slightly attenuated. However, further restricting analyses to a healthy subgroup attenuated the association for reallocating time from sedentary behaviour to light physical activity. Other associations, including those for reallocating time to MVPA, were attenuated slightly or not at all. Residual confounding also remains possible, although sensitivity analyses using a negative control outcome and E-values suggested its impact is likely to be limited. While results are presented for reallocations of time between behaviours, these are derived statistically across participants: each participant had a single measurement, so within-participant changes cannot be addressed directly. Validation of the machine-learning methods on another independent dataset would help to further understand their robustness. Finally, UK Biobank is not a population-representative cohort^15^ (e.g. low socio-economic status individuals are under-represented compared to the national population^31^), though a previous study showed exposure-outcome associations found in UK Biobank were similar to results in more representative samples.^32^

The use of machine learning and Compositional Data Analysis methods can enhance epidemiological studies using wearable device data, leading to new health insights. The results of this study support the framing of current guidelines and interventions around increasing time spent in MVPA, and reallocating time from sedentary behaviour to light physical activity where that is infeasible.^33,34^ To strengthen the evidence for causality, intervention studies should examine the health consequences of reallocating time between movement behaviours.

## Supporting information

STROBE_checklist

## Data Availability

UK Biobank data is available to researchers who submit an application (https://www.ukbiobank.ac.uk/). The summary physical activity, sedentary behaviour, and sleep variables that we have developed will be made available as a part of the UK Biobank Returns Catalogue (https://biobank.ndph.ox.ac.uk/showcase/docs.cgi?id=1). Code for accelerometer data processing, feature extraction, machine-learning, and Compositional Data Analysis is available at https://github.com/activityMonitoring.

https://github.com/activityMonitoring

## Data sharing

UK Biobank data is available to researchers who submit an application (https://www.ukbiobank.ac.uk/). The summary physical activity, sedentary behaviour, and sleep variables that we have developed will be made available as a part of the UK Biobank Returns Catalogue (https://biobank.ndph.ox.ac.uk/showcase/docs.cgi?id=1). Code for accelerometer data processing, feature extraction, machine learning, and Compositional Data Analysis is available at https://github.com/activityMonitoring.

## Author contributions

DB, AD, RW, RR, TD, KSB, MW, and KR conceptualised the study. RW, DB, and AD designed the analysis. SC, RW, and AD developed and implemented the machine-learning models for behaviour classification. RW conducted the statistical analysis, with advice from DB and AD. All authors contributed to the interpretation of results, and to the writing and revision of the manuscript. SC, RW, and AD have accessed and verified the CAPTURE-24 data. RW, SC, RR, and AD have accessed and verified the UK Biobank data.

