## Supplementary_figures for "Reallocating time from device-measured sleep, sedentary behaviour or light physical activity to moderate-to-vigorous physical activity is associated with lower cardiovascular disease risk"

**Figure S1: Participant-wise mean (a) precision and (b) recall for classification of behaviours from accelerometer data calculated in Leave-One-Participant-Out Cross-Validation (with 95% confidence interval for the mean). The x-axis gives the minimum required recorded time in the behaviour for inclusion in the calculation.**

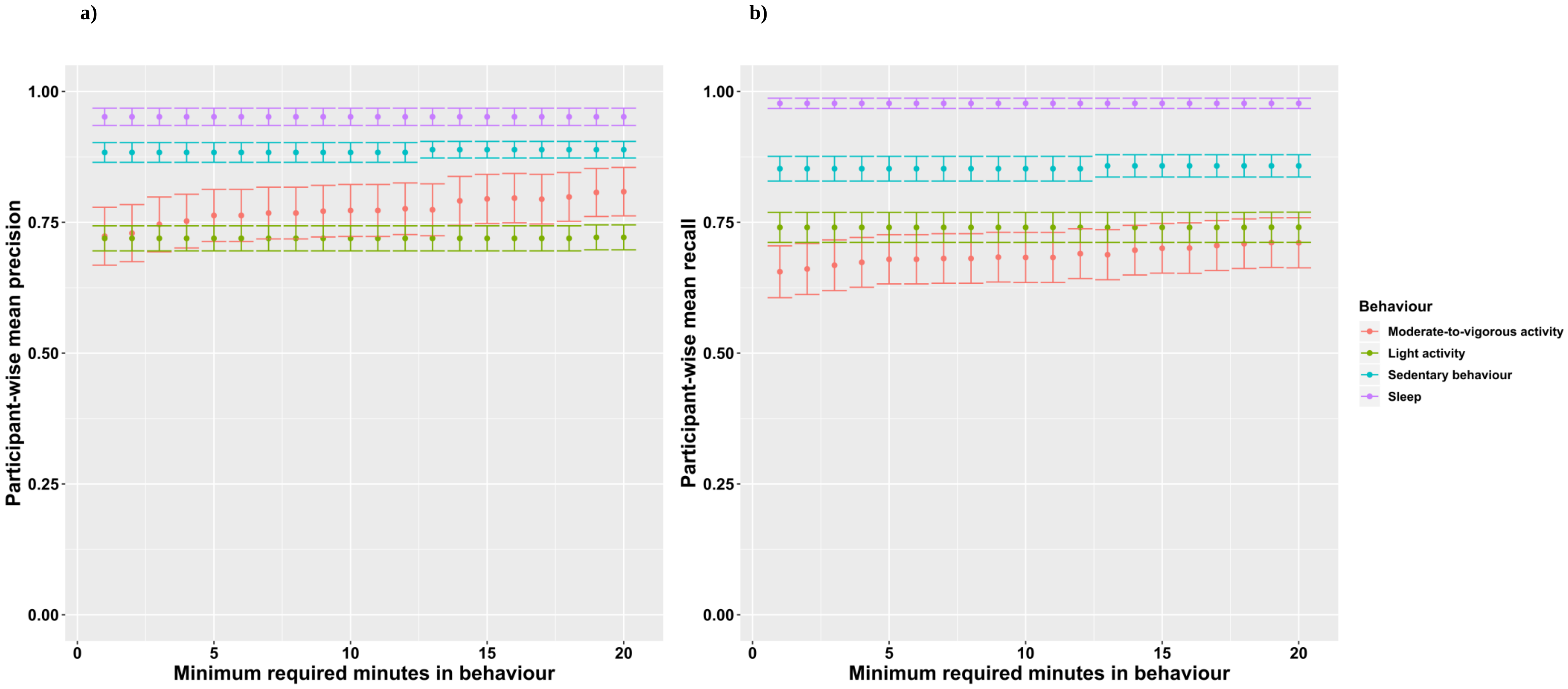

**Figure S2: Probability of being in (a) sleep, (b) sedentary behaviour (SB), (c) light physical activity (LIPA) and (d) moderate-to-vigorous physical activity (MVPA) among 87,509 UK Biobank participants according to machine-learned behaviour classification by hour of the day.**

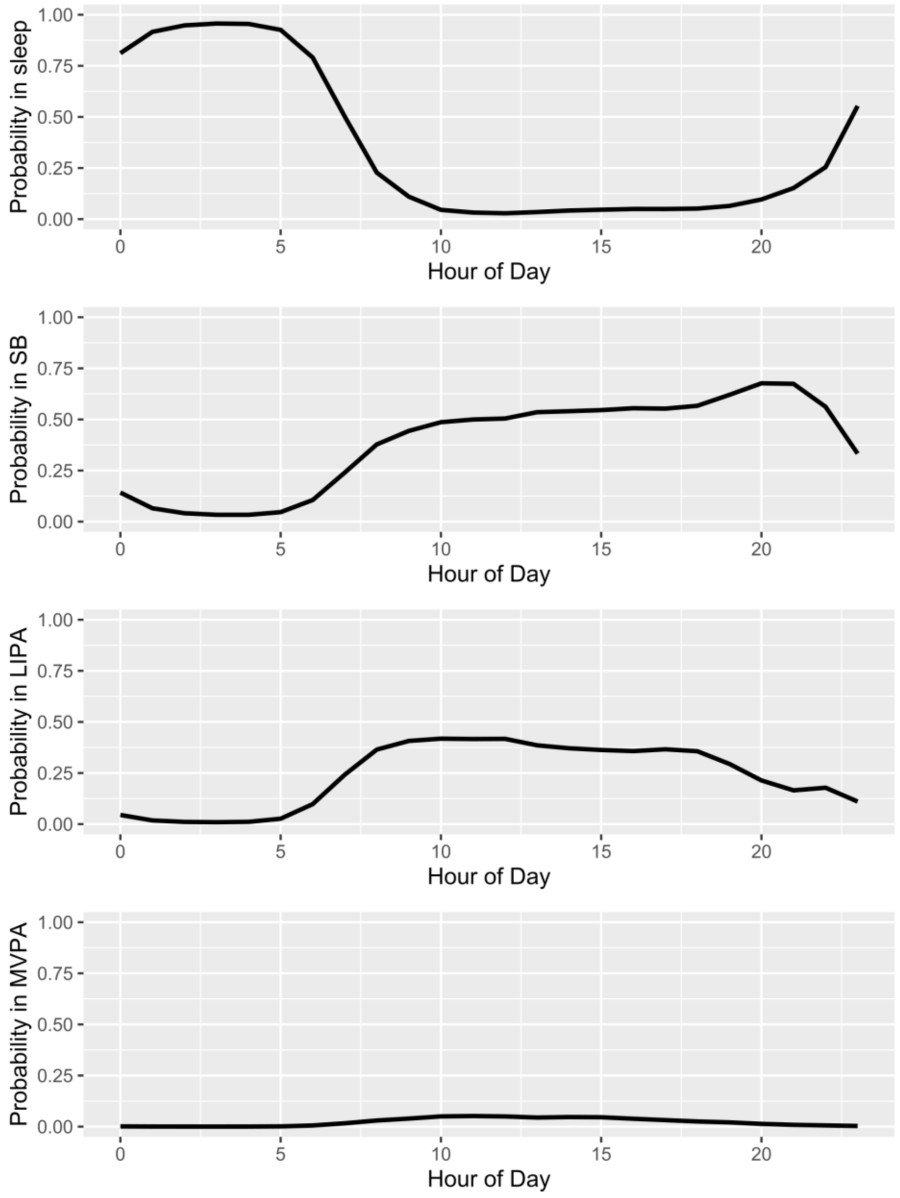

**Figure S3: Hazard Ratios for incident cardiovascular disease for all behaviour pairs estimated using a multivariable-adjusted Cox regression model .***

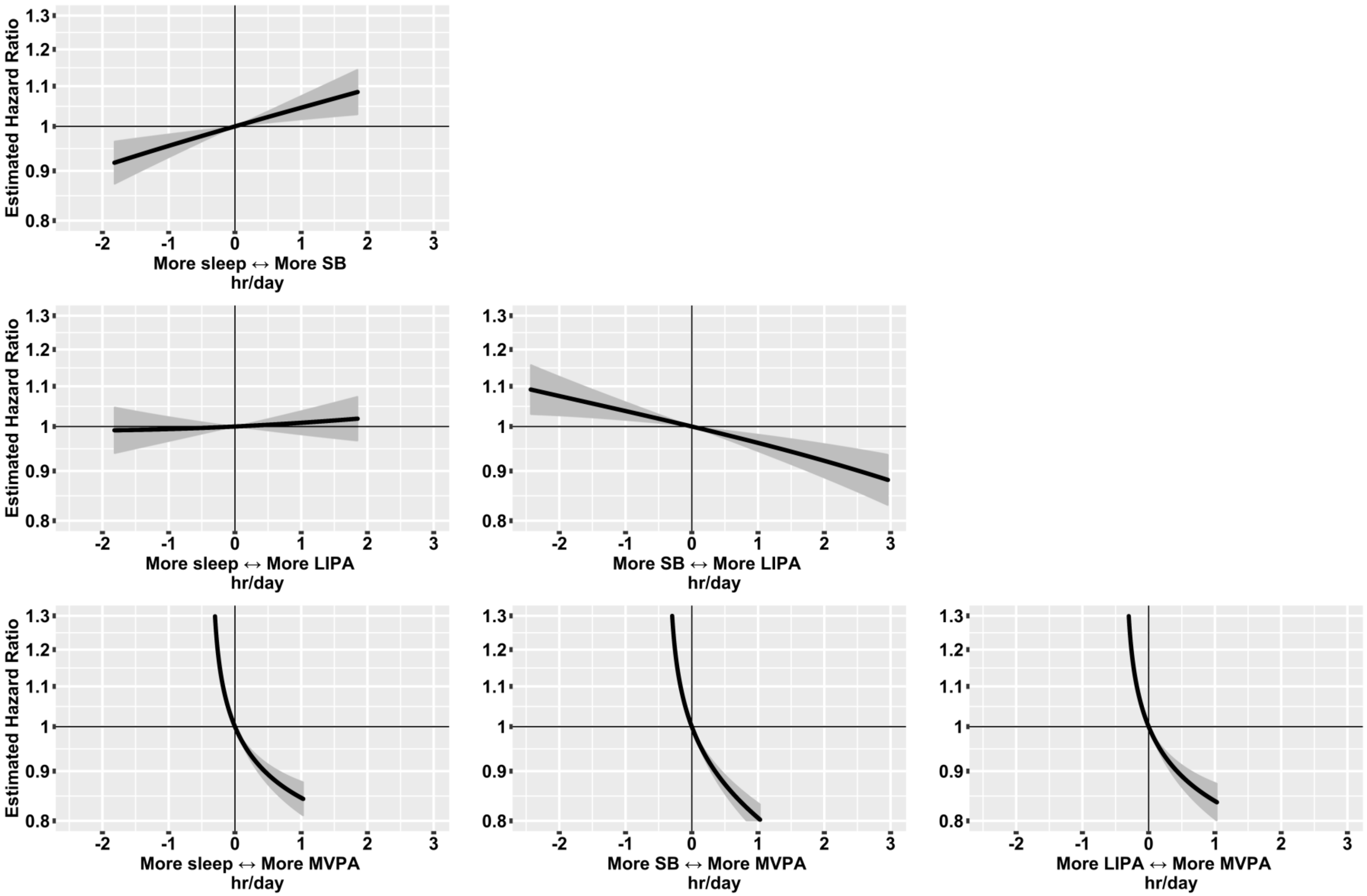


*Model based on 3,424 events in 87,509 participants. All relative to the mean behaviour composition: 8·8 hours/day sleep, 9·3 hours/day sedentary behaviour, 5·6 hours/day light physical activity, 0·35 hours/day (21 minutes/day) moderate-to-vigorous physical activity. Model used age as the timescale, was stratified by sex and was additionally adjusted for ethnicity, smoking status, alcohol consumption, fresh fruit and vegetable consumption, red and processed meat consumption, oily fish consumption, deprivation and education.

**Figure S4: Hazard Ratios for cardiovascular disease for all behaviour pairs estimated using multivariable-adjusted (blue) and minimally adjusted (red) Cox regression models .***

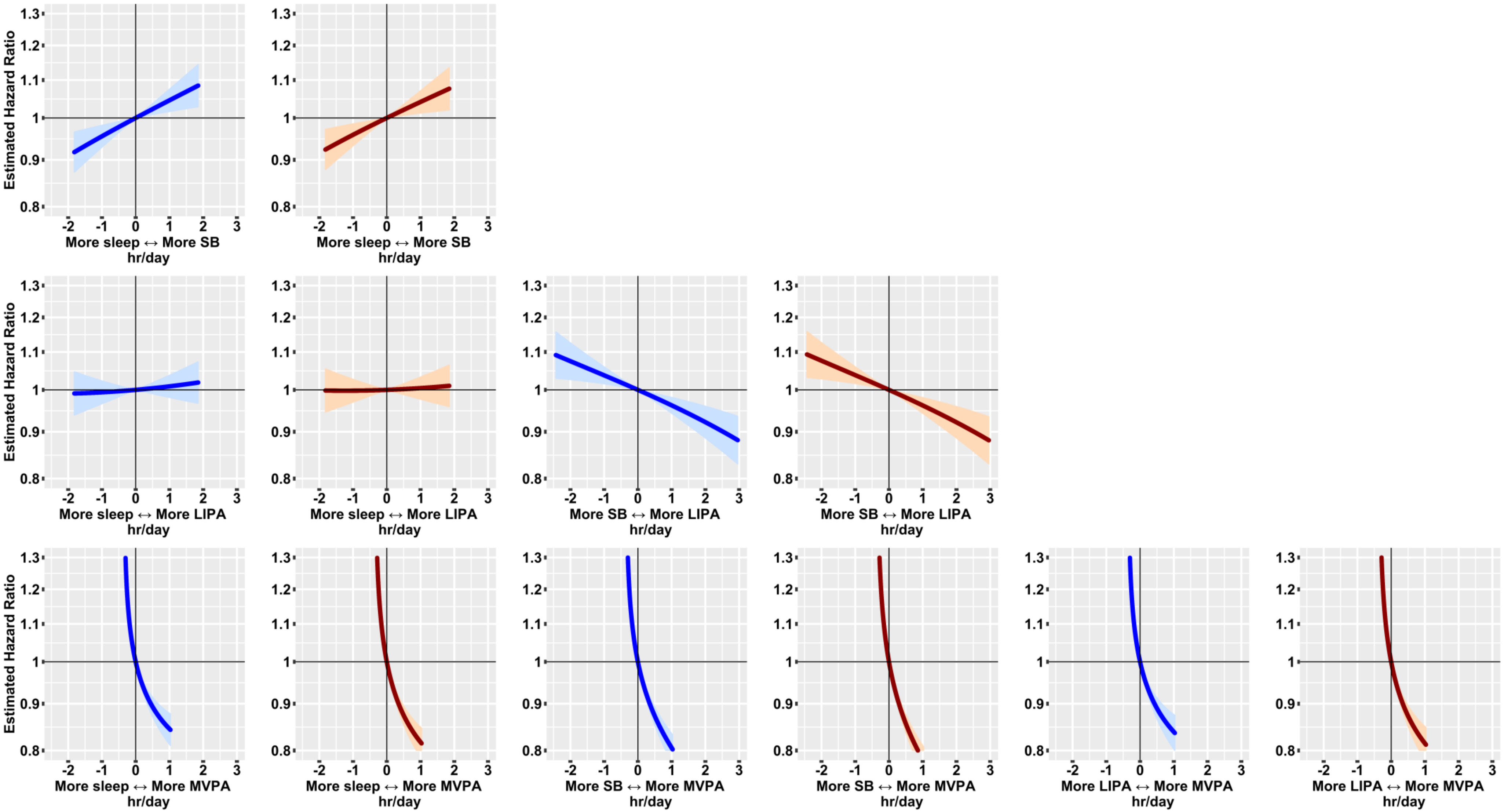


*Model based on 3,424 events in 87,509 participants. All relative to the mean behaviour composition: 8·8 hours/day sleep, 9·3 hours/day sedentary behaviour, 5·6 hours/day light physical activity, 0·35 hours/day (21 minutes/day) moderate-to-vigorous physical activity.

**Figure S5: Hazard Ratios for incident cardiovascular disease estimated using a multivariable-adjusted Cox regression model before (blue) and after (red) adjustment for BMI .***

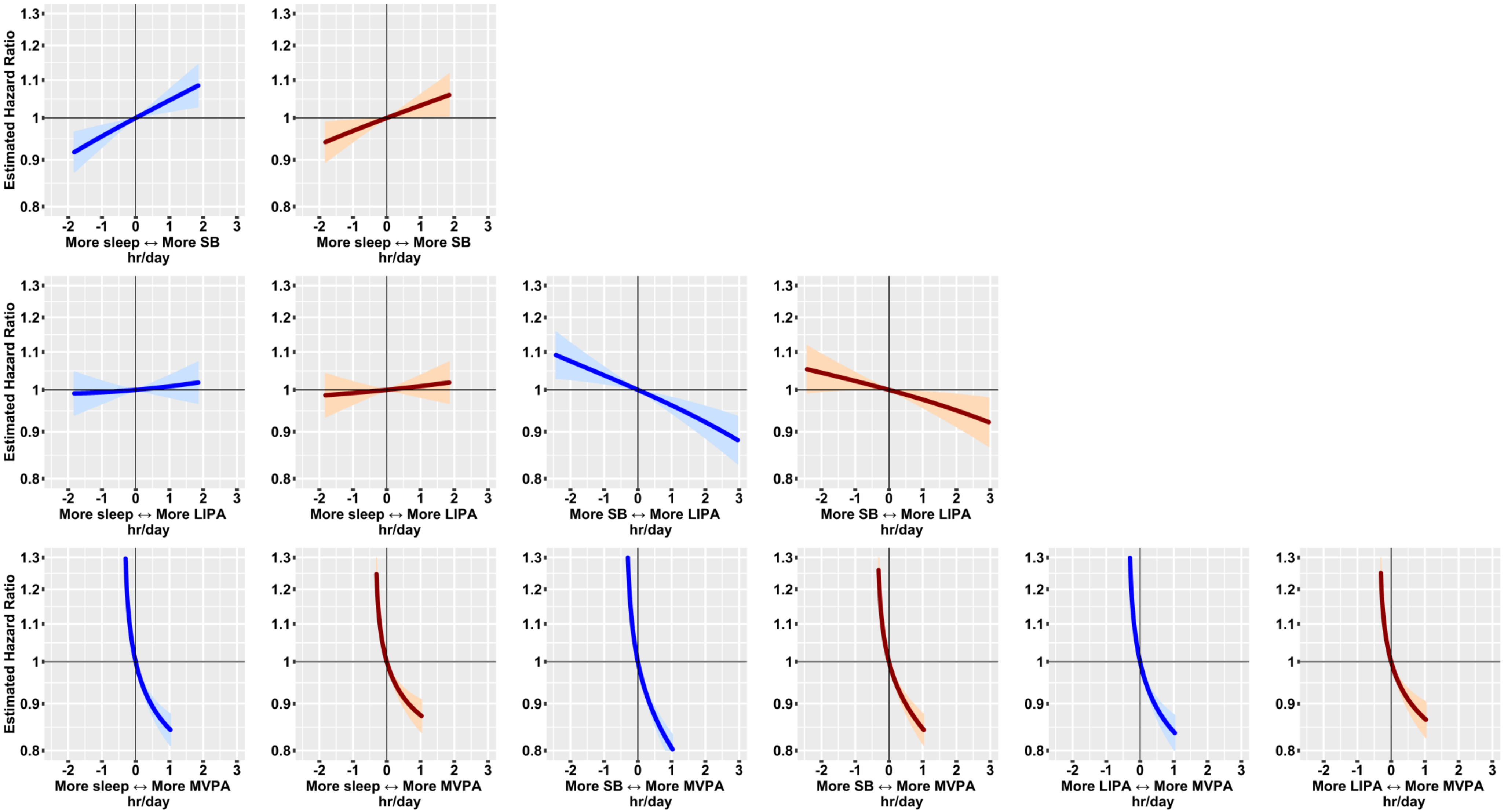


*Model based on 3,424 events in 87,509 participants. All relative to the mean behaviour composition: 8·8 hours/day sleep, 9·3 hours/day sedentary behaviour, 5·6 hours/day light physical activity, 0·35 hours/day (21 minutes/day) moderate-to-vigorous physical activity. Models used age as the timescale, were stratified by sex and were additionally adjusted for ethnicity, smoking status, alcohol consumption, fresh fruit and vegetable consumption, red and processed meat consumption, oily fish consumption, deprivation and education.

**Figure S6: Hazard Ratios for all (blue) and fatal (red) incident cardiovascular disease for all behaviour pairs estimated using a multivariable-adjusted Cox regression model .***

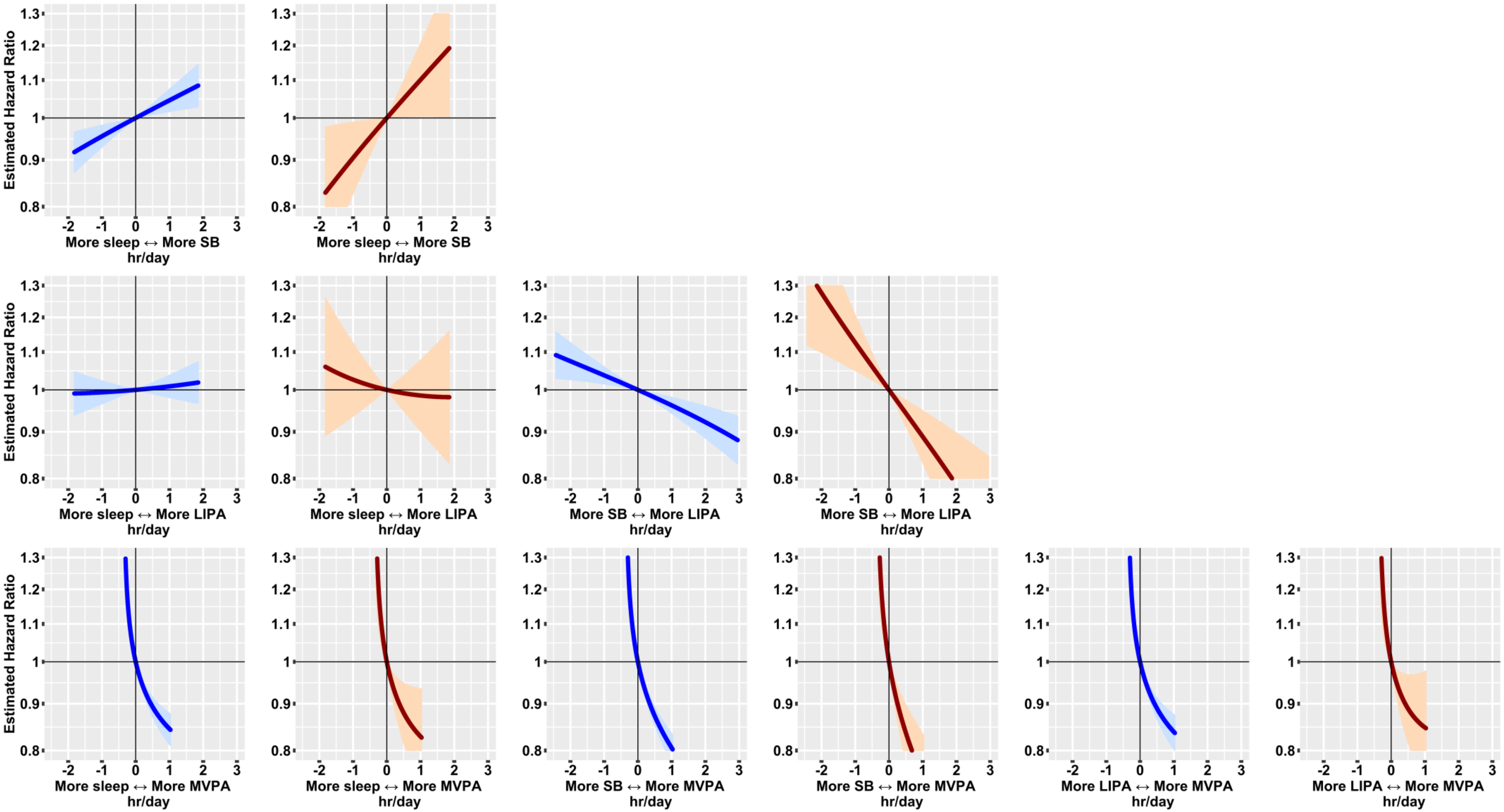


*Models based on 3,424 events and 292 deaths in 87,509 participants. All relative to the mean behaviour composition: 8·8 hours/day sleep, 9·3 hours/day sedentary behaviour, 5·6 hours/day light physical activity, 0·35 hours/day (21 minutes/day) moderate-to-vigorous physical activity. Models used age as the timescale, were stratified by sex and were additionally adjusted for ethnicity, smoking status, alcohol consumption, fresh fruit and vegetable consumption, red and processed meat consumption, oily fish consumption, deprivation and education.

**Figure S7: Hazard Ratios for incident cardiovascular disease for all behaviour pairs estimated using a multivariable-adjusted Cox regression model (blue), after removing the first two years of follow-up (red) and after additionally restricting to a healthy subgroup (green) .***

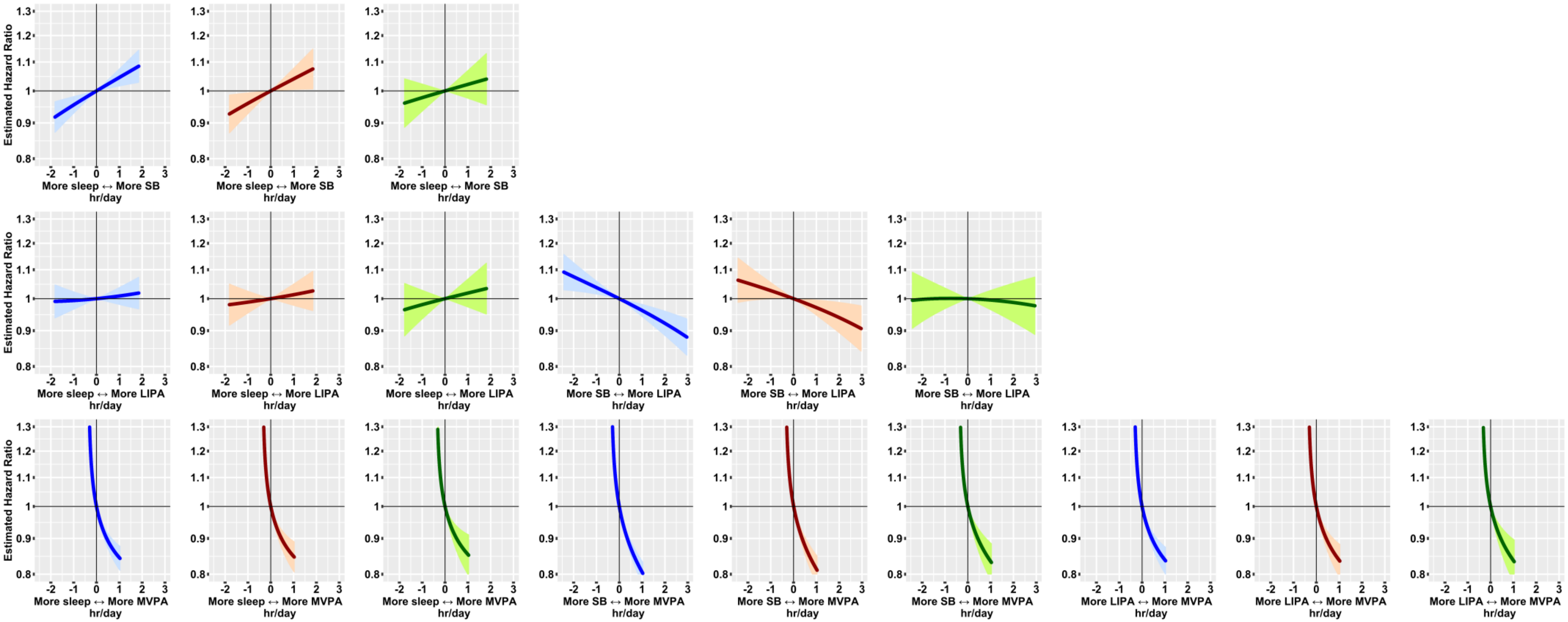


*Main analysis based on 3,424 events in 87,509 participants. First sensitivity analysis based on 2,263 events in 80,998 participants. Second sensitivity analysis based on 1,443 events in 66,075 participants. Models used age as the timescale, were stratified by sex and were additionally adjusted for ethnicity, smoking status, alcohol consumption, fresh fruit and vegetable consumption, red and processed meat consumption, oily fish consumption, deprivation and education. All values reported relative to the mean behaviour composition in each case:

Main analysis - 8·8 hours/day sleep, 9·3 hours/day sedentary behaviour, 5·6 hours/day light physical activity, 0·35 hours/day (21 minutes/day) moderate-to-vigorous physical activity

1st sensitivity - 8·8 hours/day sleep, 9·3 hours/day sedentary behaviour, 5·6 hours/day light physical activity, 0·35 hours/day (21 minutes/day) moderate-to-vigorous physical activity

2nd sensitivity - 8·8 hours/day sleep, 9·2 hours/day sedentary behaviour, 5·6 hours/day light physical activity, 0·37 hours/day (22 minutes/day) moderate-to-vigorous physical activity.

**Figure S8: Hazard Ratios for cardiovascular disease (blue) and for non-activity-related accidents (red) for all behaviour pairs estimated using a multivariable-adjusted Cox regression model .***

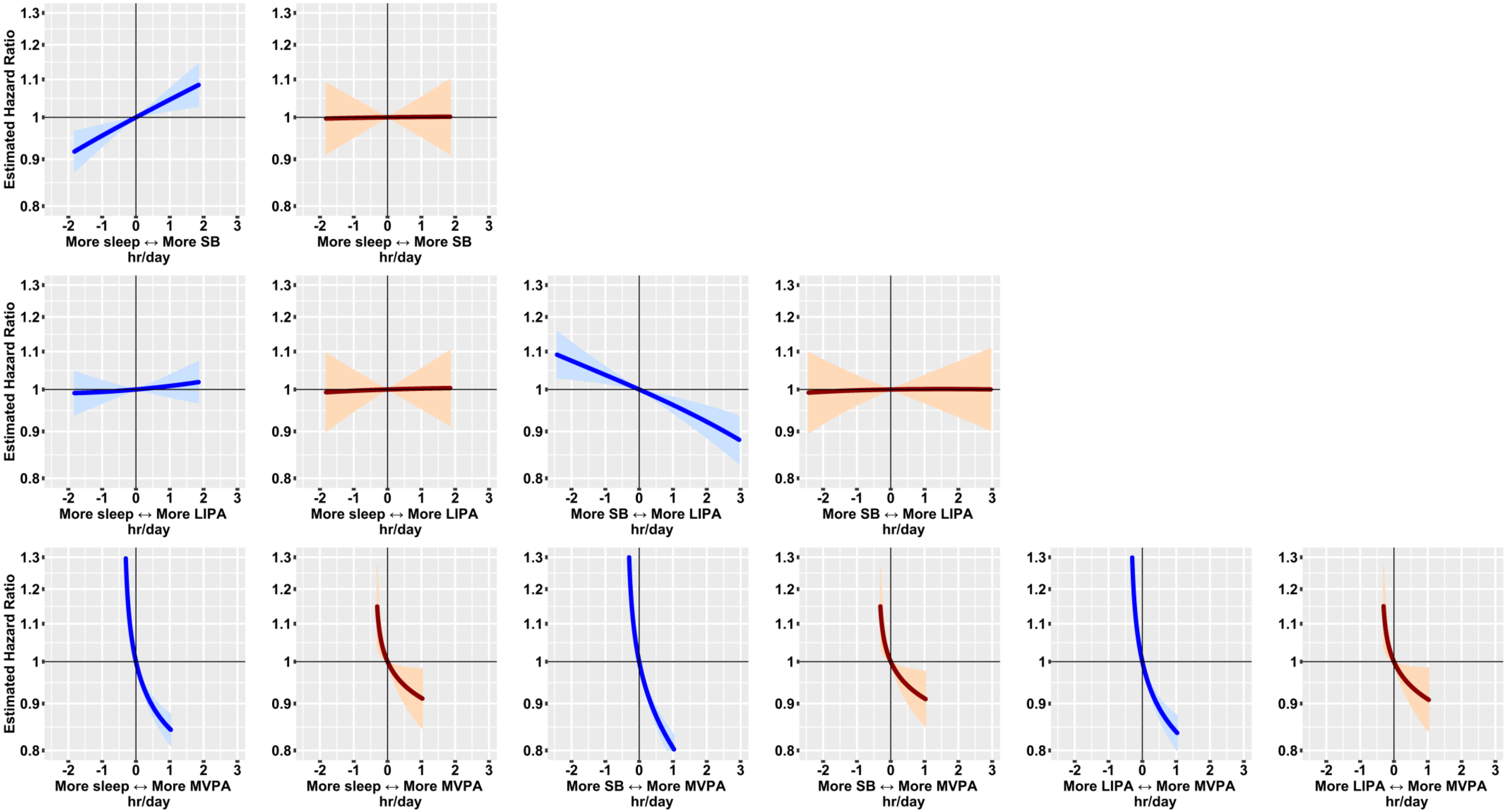


*Negative control model based on 1,187 events in 87,509 participants. All relative to the mean behaviour composition (8·8 hours/day sleep, 9·3 hours/day sedentary behaviour, 5·6 hours/day light physical activity, 0·35 hours/day (21 minutes/day) moderate-to-vigorous physical activity). Models used age as the timescale, were stratified by sex and were additionally adjusted for ethnicity, smoking status, alcohol consumption, fresh fruit and vegetable consumption, red and processed meat consumption, oily fish consumption, deprivation and education.

**Figure S9: Hazard ratios and corresponding E-values for incident cardiovascular disease associated with reallocating time to named behaviour, from all other behaviours proportionally, in 87,509 UK Biobank participants .***

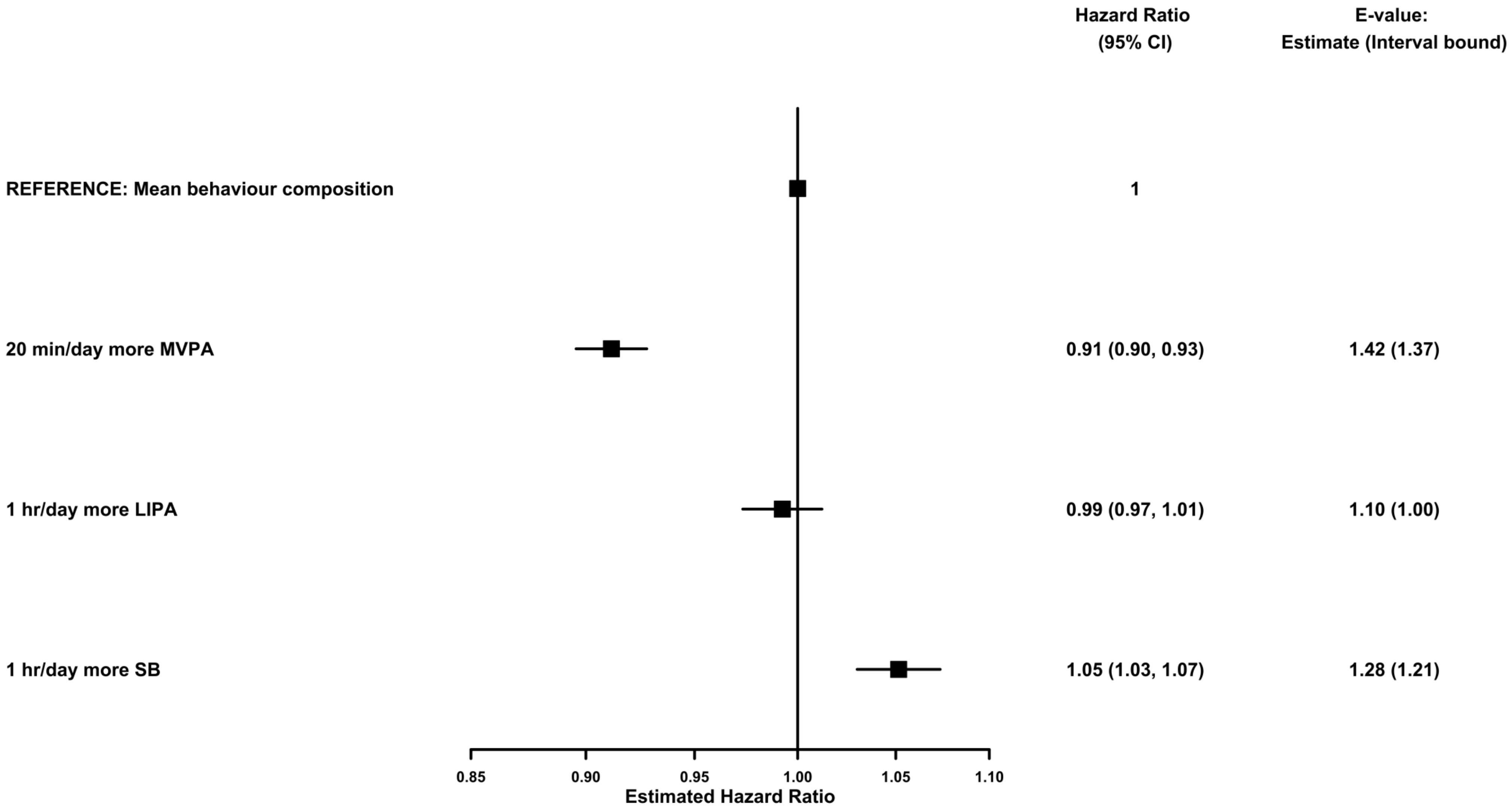


*Model based on 3,424 events in 87,509 participants. All relative to the mean behaviour composition (8·8 hours/day sleep, 9·3 hours/day sedentary behaviour, 5·6 hours/day light physical activity, 0·35 hours/day (21 minutes/day) moderate-to-vigorous physical activity) and more time in named behaviour reallocated from all other behaviours proportionally. Model used age as the timescale, was stratified by sex and was additionally adjusted for ethnicity, smoking status, alcohol consumption, fresh fruit and vegetable consumption, red and processed meat consumption, oily fish consumption, deprivation and education.

**Figure S10: An example of a decision tree to classify time windows using average acceleration vector magnitude (avm) and the 75$^{th}$ percentile of acceleration vector magnitude (75thp).**

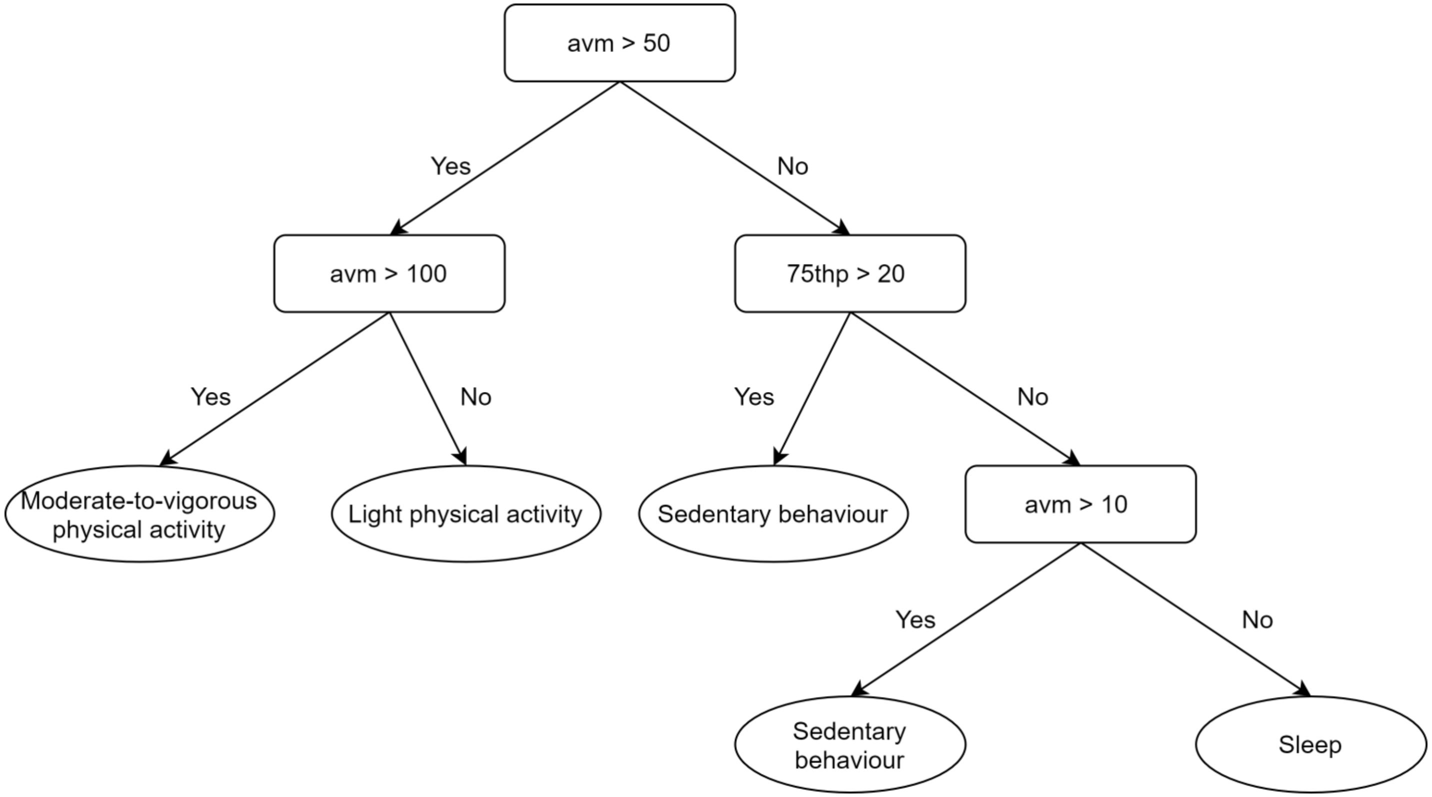

**Figure S11: The structure of a Hidden Markov Model.**

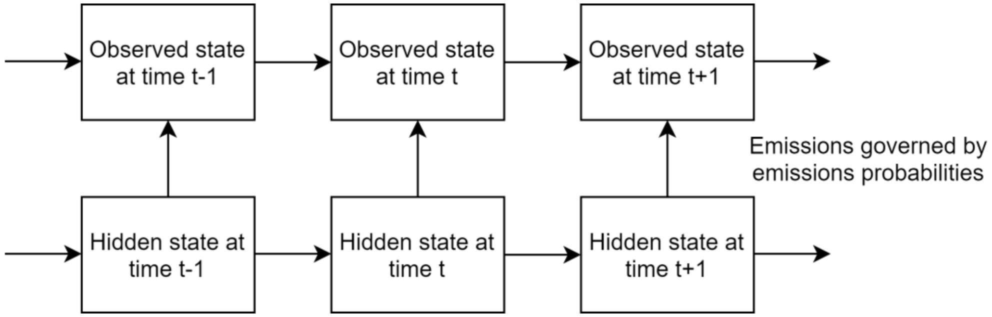

**Figure S12: Hazard Ratios for incident cardiovascular disease for all behaviour pairs estimated using a multivariable-adjusted Cox regression model for all participants (blue) and in a sensitivity analysis excluding individuals recording a zero value in any behaviour variable (red) .***

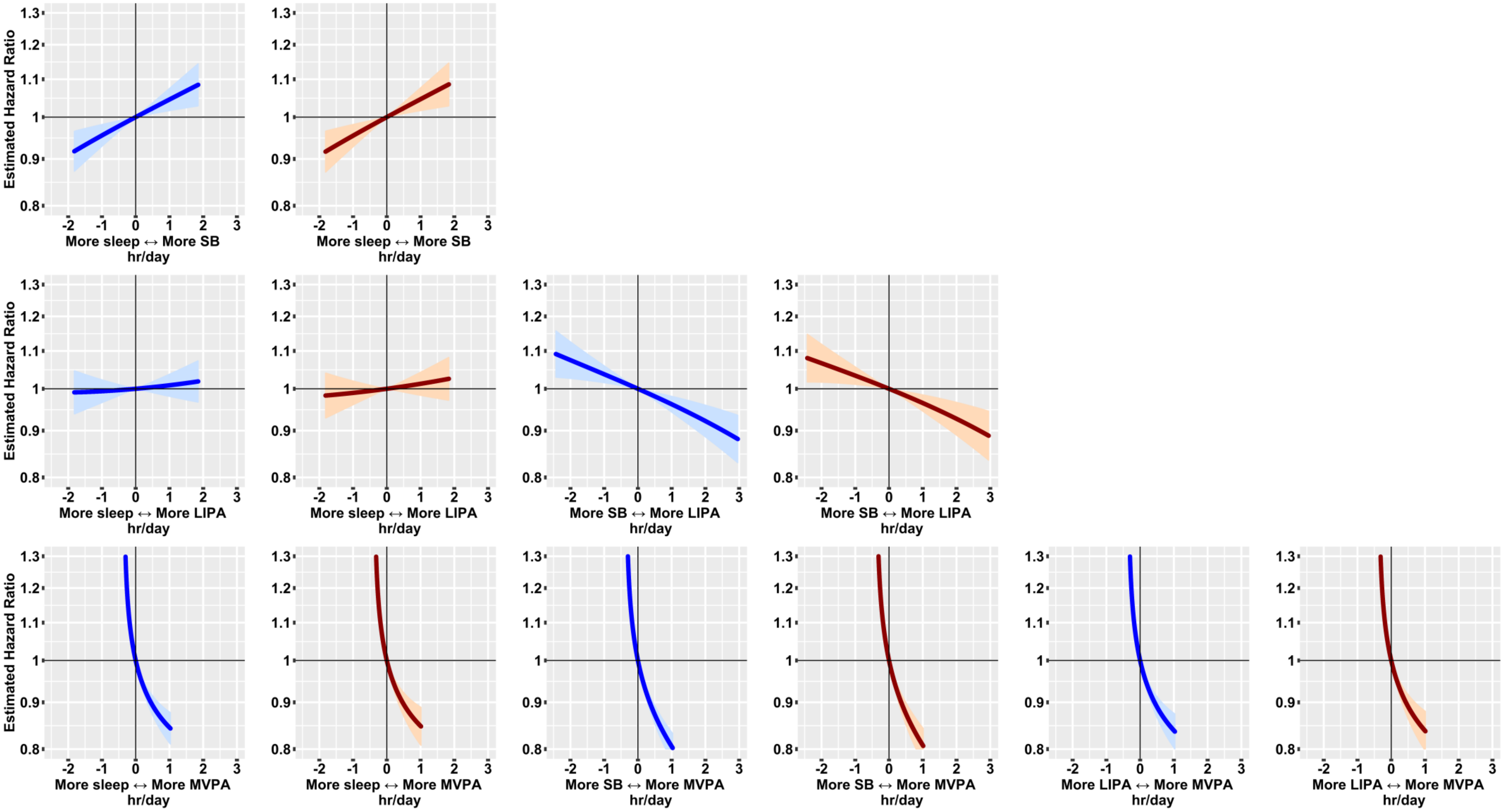


*Main model based on 3,424 events in 87,509 participants. Model excluding individuals with zero values based on 3,350 events in 86,708 participants. All relative to the mean behaviour composition in each case (main analysis – 8·8 hours/day sleep, 9·3 hours/day sedentary behaviour, 5·6 hours/day light physical activity, 0·35 hours/day (21 minutes/day) moderate-to-vigorous physical activity; analysis excluding individuals with zero values – 8·8 hours/day sleep, 9·3 hours/day sedentary behaviour, 5·6 hours/day light physical activity, 0·37 hours/day (22 minutes/day) moderate-to-vigorous physical activity). Models used age as the timescale, were stratified by sex and were additionally adjusted for ethnicity, smoking status, alcohol consumption, fresh fruit and vegetable consumption, red and processed meat consumption, oily fish consumption, deprivation and education.

**Figure S13: Hazard Ratios for incident cardiovascular disease estimated using a multivariable-adjusted Compositional Data Analysis Cox regression model (blue) and using a multivariable-adjusted linear isotemporal substitution Cox regression model (red).***

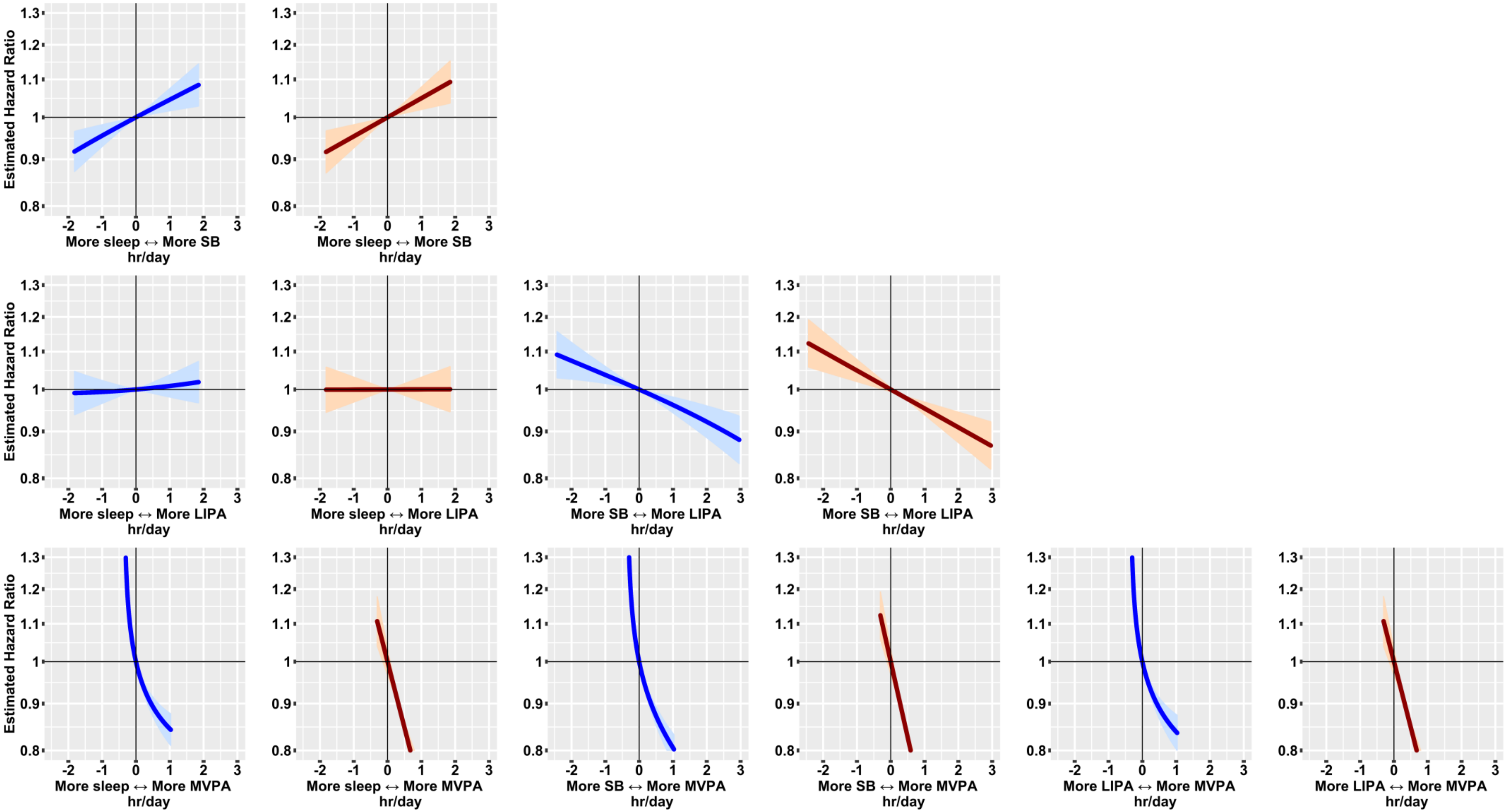


*Model based on 3,424 events in 87,509 participants. All relative to the mean behaviour composition: 8·8 hours/day sleep, 9·3 hours/day sedentary behaviour, 5·6 hours/day light physical activity, 0·35 hours/day (21 minutes/day) moderate-to-vigorous physical activity. Models used age as the timescale, were stratified by sex and were additionally adjusted for ethnicity, smoking status, alcohol consumption, fresh fruit and vegetable consumption, red and processed meat consumption, oily fish consumption, deprivation and education.
