## Supplementary_material for "Reallocating time from device-measured sleep, sedentary behaviour or light physical activity to moderate-to-vigorous physical activity is associated with lower cardiovascular disease risk"

### Search

The search, which was performed on PubMed, aimed at identifying studies on the association between device-measured movement behaviours and incident cardiovascular disease events. The search terms were “(accel*[Text Word] OR actigraph*[Text Word] OR device[Text Word] OR sensor[Text Word] OR "activity monitor"[Text Word]) AND ("physical activity"[Title] OR "sedentary"[Title] OR "sleep"[Title] OR "physical behav*"[Title] OR "time use"[Title] OR "light activity"[Title] OR "moderate activity"[Title] OR "vigorous activity"[Title] OR "moderate-to-vigorous activity"[Title] OR MVPA[Title] OR LIPA[Title] OR sitting[Title]) AND (cardiovascular[Title] OR "heart disease"[Title] OR stroke[Title] OR cerebrovascular[Title] OR "heart attack"[Title] OR "myocardial infarction"[Title])”. The search included studies published up to the end of September 2020.

Four studies were identified meeting the inclusion criteria.[1–4]

**‘Ground truth’ labelling of movement behaviours in image data**

As described in the main text, to provide the ‘ground truth’ labels for machine-learning based behaviour classification, fine-grained behaviour annotations of image data were mapped to sleep, sedentary behaviour, light physical activity and moderate-to-vigorous physical activity based on the definition in the main text. In practice, this involved the following steps (the final mapping is given in **Table S1**):

1. The fine-grained annotation for sleeping was assigned to sleep.
2. Behaviours at 3 or more METs (Metabolic Equivalent of Task, where 1 MET is energy expenditure in quiet sitting), as described in Compendium of Physical Activities,[5] were assigned to moderate-to-vigorous physical activity.
3. For waking behaviours at <3 METs, if the fine-grained annotation indicated a sitting, lying or reclining posture, the behaviour was assigned to sedentary behaviour.
4. Waking behaviours at <3 METs not assigned to sedentary behaviour were assigned to light physical activity.
5. All labels were reviewed by two reviewers. Where reviewers agreed the fine-grained annotation was typically used by annotators for behaviours in a different category to the label given, the fine-grained annotation was recoded. This review was performed prior to model training, and no changes were made after results were obtained.

### Machine-learning methods

As described in the main text, a Random Forest (RF) with 100 decision trees was developed to classify 30-second time windows as sleep, sedentary behaviour, light physical activity or moderate-to-vigorous physical activity using the time and frequency domain features outlined in **Table S2**. A Hidden Markov model (HMM) was then employed to use time sequence information to improve on the RF-assigned label sequence. As described in the main text, models were trained using labelled data from the CAPTURE-24 study, in which participants wore wearable cameras and kept time use diaries alongside wearing an accelerometer.

#### Machine-learning methods: Features

Time windows of acceleration were classified using a list of features (variables) based on features used in the study of Willetts et al (this included time and frequency domain features e.g. mean and kurtosis of the acceleration vector magnitude, and power in 1Hz bands from the Fast Fourier Transform of acceleration vector magnitude).[6] In the present study, only rotation-invariant features were used (see **Table S2**). This addressed concerns about risk of overfitting and possible time trends in the data driven by sensitivity to device orientation within the wrist strap (orientation became more standardised over 2013-2015).

#### Machine-learning methods: Random Forest models

Random Forests are based on decision trees. Decision trees assign class labels based on splits of the data using feature value thresholds (as shown in the example in **Figure S10**). They can be trained using the Classification and Regression Tree (CART) algorithm.[7]

In a RF, many decision trees are used. When training the trees (using the CART algorithm), randomness is introduced by (i) training each tree on a set of data points picked randomly (with replacement) and (ii) at each split node, restricting the choice of splitting feature to a randomly picked subset of features.

To classify a data point using an RF, it is classified by each decision tree. Because they are trained on different subsets of data and use different features, different trees in the RF may classify data points differently. The overall classification given by the RF is the class that is assigned to the data point by the largest number of trees. This approach, whereby multiple randomly-differing instances are used in order to reduce variance on the output, is the technique of bootstrap-aggregating or ‘bagging’.[8]

For this application, a balanced RF was used. The fact some behaviours are much more common than others in the labelled data (e.g. sleep is much more common than moderate-to-vigorous physical activity) can cause a standard RF, which is trained by picking N examples at random with replacement, to favour assigning common labels at the expense of less common behaviours.[6] Using the balanced RF, if there were $n_{rare}$ examples of the rarest behaviour, $n_{rare}$ examples of each behaviour were picked with replacement to train each tree.

#### Machine-learning methods: Hidden Markov models

In a HMM, there is a sequence of unobserved hidden states, which is assumed to have the Markov property (i.e. future states only depend on past states through present states). This sequence is governed by transition probabilities, which determine the probability of transitioning between each pair of states. There is a sequence of observed states, which depend probabilistically on the sequence of hidden states (described as ‘emissions’ from the sequence of hidden states; **Figure S11**).

Here, the hidden states were the true behaviours, and the emissions were the RF-assigned labels. The Viterbi algorithm, the standard approach to this problem, was used to estimate the most likely true behaviour sequence given the observed sequence of RF-assigned labels.[9] Applying the Viterbi algorithm required estimates of:

1. **Transition probabilities between hidden states:** Transition probabilities between behaviour pairs were estimated using the proportions of transitions that occurred between each behaviour pair in the labelled data.

2. **Emission probabilities of observed states from hidden states:** To estimate emission probabilities, time windows were first classified using out-of-bag predictions from the RF i.e. trees were used to classify data points on which they were not trained. This mimics use on unseen data, without requiring additional

data. Emission probabilities were then estimated using the proportions of different pairs of true behaviour and RF out-of-bag estimate.

By using this HMM to estimate the most likely true behaviour sequence given the RF-assigned labels, a more plausible sequence of states was obtained. The HMM re-labelled behaviours which formed unrealistic sequences and were likely to be attributable to misclassification (e.g. short periods of moderate-to-vigorous physical activity during sleep time). Therefore, compared to the unadjusted RF-assigned labels, the labels after using the HMM gave improved measures of the behaviours of interest for subsequent epidemiological analyses.

**Machine-learning methods: Evaluation**

All metrics were calculated in Leave-One-Participant-Out Cross-Validation.

Leave-One-Participant-Out Cross-Validation involves, for each participant, a model trained on all other participants' data (i.e. with this participant's data left-out). The trained model is then used to label the left-out participant's data and evaluation metrics are calculated. This is repeated for all participants, and metrics are aggregated or calculated across all participants.

Leave-One-Participant-Out Cross-Validation allows evaluation of the performance of the models on data not used in training, while retaining the maximal amount of data for use in training these models. Moreover, all of the data can then be used to train the final model used for classification.

For model performance, the following evaluation metrics were used:

1. We reported mean per-participant accuracy across all behaviours. This is a simple, intuitive metric of model performance, describing the proportion of 30-second time windows that were correctly classified. Using mean per-participant accuracy, rather than aggregate accuracy over all data, prevents the result being dominated by performance on a few participants with larger amounts of data (important as there may be inter-individual differences in classification performance).
2. We reported mean per-participant Cohen's kappa across all behaviours. This is a metric of interrater reliability. It evaluates how much higher the agreement between two raters (here, annotator-assigned 'ground truth' label and model-assigned label) is than that which would be achieved by chance, given the proportions in each class. It is preferable to accuracy, as it takes into account the proportions in each class (in particular, in data where some classes are dominant, a classifier assigning solely to the dominant classes can achieve high accuracy but not high Cohen's kappa).
3. We reported mean per-participant precision and recall for each behaviour. Precision for a given behaviour is the proportion of examples labelled by the model as that behaviour which are 'true' examples of that behaviour. Recall for a given behaviour is the proportion of 'true' examples of that behaviour labelled as that behaviour. Again, taking the mean across participants prevents performance being dominated by performance on participants with larger amounts of data. However, it also upweights the contribution of individuals with very small amounts of data for a given behaviour. Therefore, precision and recall were additionally calculated after excluding participants with up to 20 minutes in the behaviour.

Taken together, these metrics help to understand the validity of the model as a method derive measures of movement behaviours for subsequent epidemiological analyses. After applying the model to derive measures of movement behaviours for UK Biobank participants, face validity was assessed by plotting behaviour profiles over the day.

### A Compositional Data Analysis approach to movement behaviour data

#### Log-ratio transformation

A Compositional Data Analysis approach is a set of methods for working with compositional data, based on the use of log-transformed ratios to describe the data.[10–12] Ratios between behaviours are used to describe compositional data as they capture the relative values of the different behaviours. Log-transforming ratios ensures the relationships and distances between different compositions are well-described (using log-transformed ratios is equivalent to working with compositional data in a 'natural' space for it, with operations which map compositions to genuine compositions and an appropriate distance metric[13,14]). For statistical purposes, log-transformed ratios are also typically more conveniently distributed than ratios.[15]

While many different sets of log-transformed ratios can be used, isometric log-ratio pivot coordinates are widely used in movement behaviour research[16] and were used in this study. They were calculated as follows:

$$coordinate_1 = \sqrt{\frac{3}{4}}\ln\left(\frac{\text{sleep}}{\sqrt[3]{\text{SB}\times\text{LIPA}\times\text{MVPA}}}\right) = \sqrt{\frac{1}{12}}\ln\left(\frac{sleep}{SB}\right) + \sqrt{\frac{1}{12}}\ln\left(\frac{sleep}{LIPA}\right) + \sqrt{\frac{1}{12}}\ln\left(\frac{sleep}{MVPA}\right)$$

$$coordinate_2 = \sqrt{\frac{2}{3}}\ln\left(\frac{\text{SB}}{\sqrt[2]{\text{LIPA}\times\text{MVPA}}}\right) = \sqrt{\frac{1}{6}}\ln\left(\frac{SB}{LIPA}\right) + \sqrt{\frac{1}{6}}\ln\left(\frac{SB}{MVPA}\right)$$

$$coordinate_3 = \sqrt{\frac{1}{2}}\ln\left(\frac{\text{LIPA}}{\text{MVPA}}\right)$$

#### Interpreting isometric log-ratio pivot coordinates

As the coefficients in the model relate to the isometric log-ratio pivot coordinates, rather than the raw behaviour variables, interpreting them is not straightforward.

The first coordinate describes the balance between sleep and all other behaviours. Therefore, the coefficient of the first coordinate can be interpreted as the effect of reallocating time to sleep from all other behaviours proportionally i.e. if the coefficient of the first coordinate is greater than 0 (its exponent is greater than 1) reallocating time to sleep from all other behaviours proportionally is associated with higher risk of cardiovascular disease. However, the second and third coordinates are harder to interpret analogously. Therefore, to interpret individually the effect of reallocating time to each behaviour (from all others proportionally), and following standard methods in movement behaviour research, one model per behaviour was produced (with different first coordinate). This approach was used to present the model parameters in **Table S6** (note that, in consequence, they do not parametrise a single model).

However, even using this approach, the magnitudes of the coefficients are hard to interpret. Therefore, as described in the main text, and following established methods, model estimates of the hazard ratio at different compositions relative to the mean behaviour composition were reported e.g. using isotemporal substitution plots.

#### Zero values

All participants recorded time in sleep, sedentary behaviour and light physical activity, but 1% of participants recorded no time in moderate-to-vigorous physical activity. As zero values cannot be incorporated directly into the coordinates above, different approaches to work with them have been developed. The appropriate method depends on the source of the zero values:

1. 'Rounded' zeroes relate to measurement precision: even where no time in a given behaviour was observed, had the wear time been long enough, or the time resolution of the measurement short enough, some time in the behaviour would be expected. If data contains rounded zeroes, they can be imputed as small positive values.[17]

2. 'True' zeroes occur where no matter the precision of the measurement, no time in that behaviour would be observed. For example, this may occur in movement behaviour research if someone is physically unable to take part in certain behaviours. If data contains true zeroes, participants with a true zero in a particular behaviour should be excluded from the main analysis and analysed separately.

We followed established methods in movement behaviour research by considering zero values to be ‘rounded’ and imputing them using the log-ratio expectation maximization algorithm from the ‘zCompositions’ R package.[12,17–19] Sensitivity of results to the method of treating zero values (imputation or exclusion) under the Compositional Data Analysis approach was examined by performing an analysis restricted to participants with non-zero values in all behaviour variables. This did not materially impact the results (**Figure S12**).

**Sensitivity analyses: further details on E-values**

As described in the main text, E-values were reported alongside hazard ratios. The E-value for the estimate quantifies the minimum strength of association that an unmeasured confounder would need with both exposure and outcome to explain away the observed association. The E-value for the 95% confidence interval quantifies the minimum strength of association an unmeasured confounder would need with both exposure and outcome to reduce the interval to overlap the null.[20,21] As the exposure is continuous, in both cases the risk ratio would apply to hypothetical groups with either the specified behaviour composition or the reference (the mean behaviour composition).[20]

### Sensitivity analyses: linear isotemporal substitution

For comparability with previous literature, a sensitivity analysis using a linear isotemporal substitution approach was conducted.

Under a linear isotemporal substitution approach, all but one of the movement behaviours are included in the model (so the included variables are not perfectly multicollinear). [In this study, as non-wear time was imputed all subjects had the same wear time. Therefore, a total time variable was not included, meaning the approach may be more properly called ‘leave-one-out regression’ than true linear isotemporal substitution.[12]] Associations are modelled as linear (rather than linear in the log-ratios, as under a Compositional Data Analysis approach). The coefficient of each behaviour can be interpreted in terms of replacing time in the left-out behaviour with time in that behaviour. Linear isotemporal substitution has been widely used in movement behaviour epidemiology,[22] but has been criticised for not addressing the fact movement behaviour data only conveys relative information.[12]

While there were some differences in shape of the associations observed (due to the different assumptions), results using this approach were broadly similar to the results of the main analysis using Compositional Data Analysis isotemporal substitution (**Figure S13**).

**Software**

Data preparation and development of the machine-learning models used Python 3.6.6, with the 'biobankAccelerometerAnalysis' tool[6,23,24] for preparing accelerometer data and training machine-learning models, and the 'ukb_download_and_prep_template' tool[25] for preparing covariate and outcome data (both available at https://github.com/activityMonitoring). Statistical analysis was performed in R 3.6.2[26] with 'Epi', 'zCompositions' , 'survival', 'forestplot', 'xts', 'EValue' , 'plyr', 'data.table' and 'gtools'.[17,20,27–30] The R package 'epicoda' was developed for this analysis (also available at https://github.com/activityMonitoring).[31]
